# Changes in the cerebrospinal fluid proteome precede and stratify the course of Parkinson’s Disease

**DOI:** 10.1101/2022.06.08.22276035

**Authors:** Kazuto Tsukita, Haruhi Sakamaki-Tsukita, Sergio Kaiser, Luqing Zhang, Mirko Messa, Pablo Serrano-Fernandez, Ryosuke Takahashi

**Affiliations:** Department of Neurology, Graduate School of Medicine, Kyoto University, Kyoto, Japan; Division of Sleep Medicine, Kansai Electric Power Medical Research Institute, Osaka, Japan; Advanced Comprehensive Research Organization, Teikyo University, Tokyo, Japan; Translational Medicine Department, Novartis Institutes for Biomedical Research, Basel, Switzerland; Cardiovascular and Metabolism Department, Novartis Institutes for Biomedical Research, Cambridge, USA; Neuroscience Department, Novartis Institutes for Biomedical Research, Cambridge, USA

**Keywords:** Parkinson’s disease, cerebrospinal fluid, proteomics, signature, stratification

## Abstract

The proteome encodes for various information. By quantifying 4071 human proteins in the cerebrospinal fluid using high-throughput affinity proteomics, this study aimed to extract proteomic signatures associated with Parkinson’s disease using unbiased machine-learning and to examine their impact on Parkinson’s disease course.

From the Parkinson’s Progression Markers Initiative, we first included 312 drug-naïve patients with Parkinson’s disease without *GBA1*, *LRRK2*, and *SNCA* mutations (non-genetic Parkinson’s disease) and 161 healthy controls. Differentially expressed protein analysis identified 149 proteins that were likely to be differentially expressed with a false discovery rate of ≤ 0.2. Subsequently, a logistic regression analysis with the least absolute shrinkage and selection operator created a 55-protein-based model that quantified the degree to which non-genetic Parkinson’s disease-associated cerebrospinal fluid proteomic signatures were present in each participant in the form of a score named a “non-genetic Parkinson’s disease-associated proteomic score.” This score accurately distinguished non-genetic Parkinson’s disease from healthy controls in both a derivation cohort (area under the curve = 0.91 [95% confidence interval, 0.89–0.94]) and an independent validation cohort comprising 38 drug-naïve patients with non-genetic Parkinson’s disease and 15 healthy controls (area under the curve = 0.87 [95% confidence interval, 0.76–0.99]).

Notably, this score was also significantly increased in patients with Parkinson’s disease harboring *GBA1*, *LRRK2*, or *SNCA* mutations (*n* = 258), albeit to varying degrees depending on the type of mutation. Furthermore, the score of genetic prodromal individuals (*n* = 365) was intermediate between that of healthy controls and patients with Parkinson’s disease.

Next, cross-sectional correlation analyses revealed that regardless of the presence or absence of genetic mutations, this score was significantly correlated with several baseline clinical parameters and biomarkers. Furthermore, longitudinal survival analyses revealed that even after adjustment of baseline characteristics, this score significantly predicted progression to important clinical milestones including mild cognitive impairment, dementia, and Hoehn and Yahr stage IV. Finally, longitudinal analyses using linear-mixed effects models confirmed that even after adjustment of baseline characteristics, the score was significantly associated with subsequent motor and cognitive decline.

Collectively, our study demonstrated that cerebrospinal fluid proteomic signatures associated with non-genetic Parkinson’s disease could be quantified using a novel non-genetic Parkinson’s disease-associated proteomic score. Furthermore, we identified the presence of these signatures in genetic Parkinson’s disease to varying degrees depending on the type of mutation. Moreover, these signatures appeared from the prodromal stage and were robustly linked to both subsequent motor and cognitive decline in early Parkinson’s disease, indicating that the cerebrospinal fluid proteome encodes important information for both the onset and progression of Parkinson’s disease.

## Introduction

Proteins are the principal regulator of biological phenomena. After being produced within cells, proteins enter the extracellular space by intentional secretion from the cell or by leakage due to cell damage or cell death.^1^ Therefore, the composition of extracellular proteins represents a valuable source for capturing dynamic biological phenomena occurring in the body.^1–3^ Recent advances in affinity proteomics technology have enabled the easy measurement of thousands of proteins simultaneously.^4–6^ Importantly, the reliability of this measurement method has been demonstrated to be high (intra- and inter-plate median coefficient of variation < 5%),^4, 5^ which permits the capturing of changes in the proteome with unprecedented precision. Indeed, recent large-scale landmark studies have shown that information concerning genomic and environmental factors, aging, and various health problems can be deciphered from the proteome using the affinity proteomics technology.^1, 2, 7^

Although cerebrospinal fluid (CSF) is relatively difficult to obtain in routine clinical practice, it represents an attractive resource for studying biological phenomena in the brain and identifying biomarkers for neurodegenerative diseases because brain-derived proteins are concentrated in the CSF.^8, 9^ Previous studies have reported that CSF levels of certain proteins linked to Parkinson’s disease (PD) pathogenesis including α-synuclein differ between patients with PD and healthy controls (HCs),^10–13^ and that disease-relevant forms of these proteins, such as oligomeric and phosphorylated α-synuclein, can also be quantified in the CSF.^14–16^ Recently, even α-synuclein “seeds” that induce α-synuclein aggregation can be detected in the CSF.^17, 18^ From a proteomic perspective, a recent important study measured 341 proteins in the CSF of 53 patients with PD and 72 HCs using liquid chromatography-tandem mass spectrometry, and examined the presence of PD-associated changes in the CSF proteome.^19^ However, as with another important CSF proteomic study,^20^ the sample size and/or number of proteins measured were probably too small to provide a complete picture of PD-associated changes in the CSF proteome. In addition, their impact on the course of PD remains completely unclear.

One of the popular methods for capturing disease-associated changes in the proteome (i.e., disease-associated proteomic signatures) is to aggregate information on multiple proteins in the proteome, create a model that best represents differences in proteomic characteristics of the disease compared to those of HCs, and compute a score.^1, 2, 19^ The virtue of this methodology is that this can reflect many biological processes and that each patient has a different score. Therefore, this methodology should be appropriate for highly heterogeneous diseases, such as PD, where many biological processes intersect to form a slightly different pathology in each individual.^21–25^ It should be noted that patients with PD were often subgrouped by the presence or absence of *GBA1*, *LRRK2*, and *SNCA* mutations because these mutations affect the PD phenotype and biomarker profiles,^26–32^ indicating the possibility that proteomic characteristics are also different among these subtypes. With this background, the aim of this study, which measured 4071 human proteins in the CSF of 1149 subjects recruited in the Parkinson’s Progression Markers Initiative (PPMI) study, was threefold. First, we adopted an unbiased machine-learning approach to extract CSF proteomic signatures in patients with PD without *GBA1*, *LRRK2*, and *SNCA* mutations (henceforth referred to as non-genetic PD) in the form of a score. Second, using this score, we examined whether non-genetic PD-associated CSF proteomic signatures were also present in patients with PD and prodromal individuals harboring *GBA1*, *LRRK2*, or *SNCA* mutations. Third, we examined the association of this score with baseline factors and longitudinal disease course.

## Materials and methods

### Quantification of CSF proteins using modified aptamers

Using Novartis’ proprietary platform version of SomaScan®, SomaLogic Inc. (Boulder, USA), blinded to clinical information, quantified proteins in the CSF collected at baseline visits for participants enrolled in the original PPMI study from June 2010 to May 2019. SomaScan® quantified each protein as relative fluorescence units of *slow offrate modified aptamer*s (SOMAmers) that bound to each target protein in slide-based hybridization microarrays. The details of this methodology have been extensively described previously.^4, 5^

Novartis’ proprietary platform version of SomaScan® contains 5080 fluorescently labeled SOMAmers to measure 4230 proteins. After removal of 298 SOMAmers that were used as control SOMAmers or targeted non-human proteins, 4782 SOMAmers targeting 4135 human proteins remained. Most SOMAmers have a one-to-one correspondence with their target protein, but some target the same protein, while others target two (or more) proteins simultaneously. While SOMAmers targeting more than two proteins were also included in the analysis in an article related to ours,^33^ we excluded them from the current analysis to enable future application using other platforms. Finally, in this study, we quantified the protein levels of 4071 human proteins targeted by 4716 SOMAmers.

Raw SomaScan® data were first subjected to sequential standardized data normalization procedures, namely Hybridization Control Normalization, Median Signal Normalization, Plate Scaling, and Calibration, by SomaLogic Inc. (Boulder, US).^5^ Following the vendor’s data standardization, relative fluorescent units were converted to a log2 scale and normalized to the global median (across all plates) divided by dilution level. Finally, the dataset was adjusted for batch effects using an empirical Bayesian method implemented in the “ComBat” function in the R package “sva.”^34^ The plate ID was selected as the batch variable, assuming that differences between shipments at the plate level would overlie differences between plates within a shipment (note that there were two shipments in this project, one in February 2019 and one in August 2019).

### Study participants

Participants in this study were selected from participants in the original PPMI study for whom high-throughput proteomics was performed. First, patients with non-genetic PD who did not have known high-risk variants of *GBA1*, *LRRK2*, and *SNCA* genes, and HCs were included. Subsequently, they were randomly divided into two groups at a ratio of 9 (derivation cohort): 1 (validation cohort) to extract and validate CSF proteomic signatures associated with non-genetic PD. Since patients with non-genetic PD included in this study were recruited from the original cohort of the PPMI study, they were drug-naïve, had Hoehn and Yahr (H&Y) stages I or II within 2 years of diagnosis, and had loss of presynaptic dopamine nerve endings confirmed by dopamine transporter (DAT) imaging. For additional analysis, we also included patients with PD and non-PD participants harboring known high-risk variants of *GBA1*, *LRRK2*, and *SNCA* genes (henceforth referred to as genetic PD and genetic prodromal, respectively). Since most of patients with non-genetic PD included in this study were recruited from the genetic cohort of the PPMI study, they were not necessarily drug-naïve and had H&Y stages I, II, or III within 7 years of diagnosis. The demographic, genetic, and clinical data used in this study were obtained from the PPMI database on December 07, 2021. For further details on the study design and updates, please visit www.ppmi-info.org.

### Details of the *GBA1*, *LRRK2*, and *SNCA* mutations

The presence or absence of protein-coding variants in *GBA1* and *LRRK2* genes was determined by the Genetics Coordination Core of the PPMI study using a series of genotype data including whole-exome, whole-genome, and targeted Sanger sequencing data, and the consensus *GBA1* and *LRRK2* coding variant summary is provided in the PPMI database (www.ppmi-info.org/access-data-specimens/data). Based on the recommendations for determining PPMI eligibility for the genetic cohort of the PPMI study (https://www.ppmi-info.org/study-design/research-documents-and-sops) and previous studies,^27, 35^ we determined the *GBA1* and *LRRK2* coding variants that is considered pathogenic or likely pathogenic with a moderate to high risk of PD. Specifically, for *GBA1* coding variants, p.84GG, IVS2+1G>A, p.R120W, p.S173*, p.E326K, p.T369M, p.N370S, p.E388K, p.L444P, p.R463C, and p.R496H were considered as *GBA1* mutations. For *LRRK2* coding variants, p.N1437H, p.R1441C/G/H, p.G2019S, and p.I2020T were considered as *LRRK2* mutations.^36^ It should be noticed that in an article related to ours,^33^ patients with p.E326K, p.T369M, and p.A456P variants in the *GBA1* gene were excluded because it focused on severe *GBA1* mutations. However, we included these patients in this study because we put focus on examining the utility of the ngPD-ProS in as many patients with PD as possible.

For *SNCA* genotyping, information regarding single nucleotide polymorphisms (rs104893877) was extracted from the whole-genome sequencing data to determine the presence of a pathogenic p.A53T *SNCA* mutation (*SNCA* mutation).^37–39^ Considering the substantially higher penetrance of the *SNCA* mutations compared to that of the *GBA1* and *LRRK2* mutations, patients with both *SNCA* mutations and *GBA1* or *LRRK2* mutations were classified as having *SNCA* mutations.

### Clinical evaluations, DAT imaging, CSF biomarkers, and longitudinal clinical milestones

For demographic information, the data on age, sex, race and ethnicity, disease duration since onset, H&Y stage in the off state, and levodopa-equivalent daily dose (LEDD) were collected.^40^ Race and ethnicity were self-reported by the study participants at the screening visit of the PPMI study. The off score of Movement disorder society-sponsored revision of the unified Parkinson’s disease rating scale (MDS-UPDRS) total score and MDS-UPDRS part III score were used as indicators of overall symptoms and motor symptoms, respectively.^41^ The MDS-UPDRS part I and part II scores were used as measures of overall non-motor symptoms and activities of daily living, respectively. The Montreal cognitive assessment (MOCA) total score was used as a measure of the global cognitive function.^42^ For subdomain measures of cognitive function, we used the Hopkins verbal learning test-revised (delayed recall T score) for verbal recent memory function,^43^ the judgment of line orientation (age and education-adjusted total score) for visual and spatial processing function,^44^ and the symbol digit modalities test (T score) for processing speed function.^45^ Depression symptoms, autonomic symptoms, daytime sleepiness severity, and dream-enacting behavior were assessed using the 15-item version of the geriatric depression scale (total score),^46^ scales for outcomes in PD-autonomic (total score),^47^ Epworth sleepiness scale (total score),^48^ and rapid-eye-movement sleep behavior disorder screening questionnaire (total score),^49^ respectively.

To evaluate the integrity of presynaptic dopaminergic terminals using DAT imaging, specific binding ratio (SBR) of the left and right caudate and putamen were used. DAT imaging were obtained with a Siemens or General Electric single photon emission computed tomography system 3-4 h following ^123^I-ioflupane injection. DAT imaging data were assessed by the Imaging Core of the PPMI study, and SBR of the caudate and putamen were calculated.^50^

For CSF biomarkers, we used data on amyloid-β 1 to 42, total-tau, phosphorylated tau at threonine 181, α-synuclein, and neurofilament light chain, which were measured using the high-precision Roche Elecsys electrochemiluminescence immunoassay as reported in previous studies.^13, 51^ Further details on data collection are available on the PPMI website (www.ppmi-info.org/study-design/research-documents-and-sops).

For longitudinal clinical milestones, progression to H&Y stages III and IV, and the development of mild cognitive impairment (MCI) and dementia was used to follow the severity of the motor and cognitive symptoms, respectively. The presence of MCI was determined based on the following criteria: (1) cognitive complaint by either the patient or the informant (spouse, family member or friend), (2) cognitive impairment defined as at least 2 test scores (out of 6 scores) from at least 1 domain (out of 4 domains) > 1.0 standard deviation below the standardized mean, and (3) no functional impairment as a result of cognitive impairment.^52^ The presence of dementia was determined based on the following criteria: (1) history of cognitive decline determined by the investigator based on information from the patient, other informant (spouse, family member or friend) and the investigator’s judgment, (2) cognitive impairment defined as at least 1 test score (out of 6 scores) from at least 2 domains (out of 4 domains) > 1.5 standard deviation below the standardized mean, and (3) functional limitation as a result of cognitive impairment.^53^ The evaluation of H&Y stage was basically conducted at every visit predefined in the original PPMI study. The evaluation of MCI and dementia was basically conducted annually. Results of evaluations are provided in the PPMI database (www.ppmi-info.org/access-data-specimens/data).

### Statistical analysis

All statistical analyses were conducted by K.T. using self-made R scripts. Throughout the study, multiple testing was adjusted using the Benjamini & Hochberg method,^54^ which controls the false discovery rate and computes *q* values based on individual *P* values. We used two-sample t-test and Fisher’s exact test for group comparisons, as appropriate.

Extraction of CSF proteomic signatures associated with non-genetic PD was conducted as follows. First, differentially expressed protein analysis was conducted using the linear model implemented in the R package “limma,” adjusted for age, sex, and principal components 1– 4.^33, 55^ SOMAmers with *q* values of ≤ 0.2 were defined as SOMAmers that were likely to be differentially expressed,^56, 57^ and proteins targeted by these SOMAmers were defined as proteins that were likely to be differentially expressed. Second, using these SOMAmers, we applied logistic regression with the least absolute shrinkage and selection operator (LASSO) regularization.^58^ The tuning parameter λ for the LASSO was chosen by 10-fold cross validation using the R package “glmnet.”^59^ Subsequently, the logit value computed from the tuned model was termed a “non-genetic PD-associated proteomic score (ngPD-ProS).” We adopted this two-step methodology to reduce the risk of over-fitting and to build the model with as few proteins as possible for future clinical applications.^58, 59^ Kyoto Encyclopedia of Genes and Genomes (KEGG) and Reactome pathway enrichment analyses were conducted using the R package “clusterProfiler” and “ReactomePA,”^60, 61^ respectively.

Diagnostic accuracy was evaluated using the area under the receiver operating characteristic curve (AUC) and 95% confidence intervals (CIs) were calculated using DeLong’s algorithm with the R package “pROC.”^62^ We also calculated the odds ratio (OR) using a logistic regression model adjusted for age and sex, and 95% CIs were calculated via the profile likelihood method. For cross-sectional correlation analysis adjusted for covariates, we calculated the Pearson’s partial correlation coefficient (*r_par_*) using the R package “ppcor.”^63^ For longitudinal survival analyses, using the R package “survival,” we first computed the hazard ratio (HR) adjusted for covariates via the Cox proportional hazards model. Further, we conducted Kaplan–Meier analysis with statistical significance evaluated using the log-rank (Mantel−Cox) test and 95% CIs were calculated via the pointwise CI method. In addition, using the R package “lme4,”^64^ we examined the interaction effect between non-genetic PD-associated CSF proteomic signatures and longitudinal changes in motor and cognitive symptoms using the multivariate linear mixed effects model (random intercept/slope model with parameters being estimated using maximum likelihood method).^65^ In this model, each participant identification number was included as a predictor variable with random effects. The statistical significance on adding the interaction term to the model were examined using the likelihood ratio test.

Sensitivity analysis for the longitudinal analysis on clinical milestones was performed by limiting the follow-up period to 5 years. Two-sided *P* values and *q* values ≤ 0.05 were considered statistically significant. Values are presented as mean ± standard deviation or 95% CIs.

### Data availability

All data used in this study are available for certified investigators in the PPMI database. The R scripts used in this study will be made publicly available upon acceptance of the manuscript.

## Results

### Clinical characteristics

An overview of this study is presented in Fig. 1. For the extraction of non-genetic PD-associated CSF proteomic signatures, we included 312 drug-naïve patients with non-genetic PD and 161 HCs for the derivation cohort and 38 drug-naïve patients with non-genetic PD and 15 HCs for the validation cohort. For additional analyses, we included 258 drug-naïve patients with genetic PD (genetic PD cohort) and 354 genetic prodromal participants (genetic prodromal cohort). The background characteristics of participants in the derivation and validation cohorts are summarized in Table 1. The study flowchart is presented in Supplementary Fig. 1.

**Figure 1.**
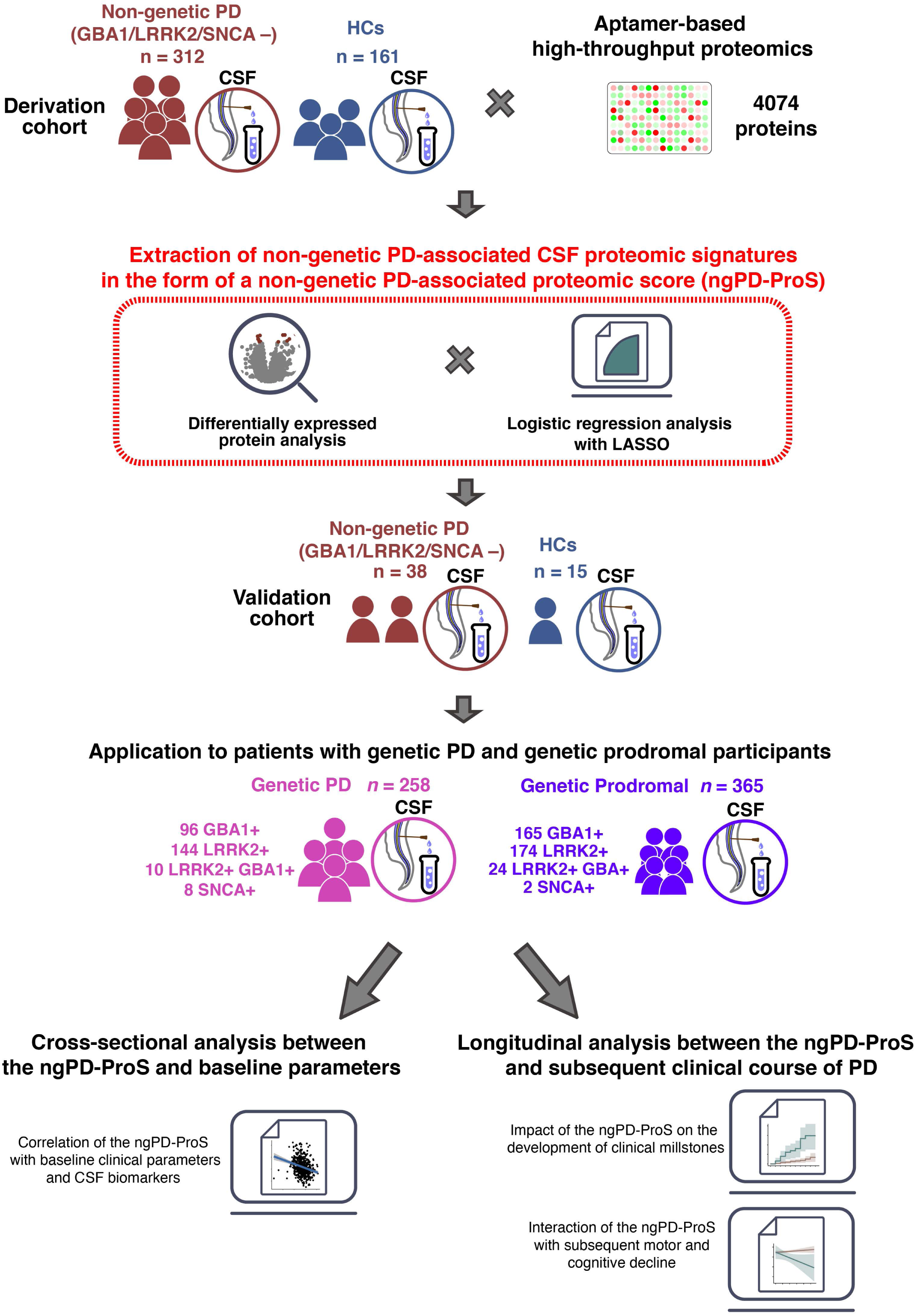
Study overview. In this study, 4074 proteins in the CSF were quantified via high-throughput affinity-based proteomics. Affinity-based proteomic data of patients with PD without *GBA1*, *LRRK2*, and *SNCA* mutations, and HCs in the derivation cohort were coupled with machine-learning to construct a “non-genetic Parkinson’s disease-associated proteomic score (ngPD-ProS).” Subsequently, the ngPD-ProS was validated in the independent validation cohort. Additionally, the ngPD-ProS was calculated for both genetic PD and genetic prodromal cohorts. Furthermore, the impact of the ngPD-ProS on the stratification of patients with PD was evaluated in a multifaceted manner through cross-sectional correlation and longitudinal analyses.

**Table 1.** Baseline characteristics of the enrolled particip ants to derive and validate the ngPD-ProS that quantified non-genetic PD-associated CSF proteome signatures.

| Characteristic | Derivation |  | Validation |  |
| --- | --- | --- | --- | --- |
|  | Non-genetic PD<br>(n = 312) | HCs<br>(n = 161) | Non-genetic PD<br>(n = 38) | HCs<br>(n = 15) |
| Age, years | 62.2 ± 9.8 | 60.6 ± 11.4 | 61.8 ± 9.2 | 60.8 ± 14.1 |
| Female | 100 (32.1%) | 60 (37.3%) | 17 (44.7%) | 5 (33.3%) |
| Duration from the diagnosis, years | 0.5 ± 0.7 | – | 0.5 ± 0.7 | – |
| Off H&Y stage | 1.6 ± 0.5 | 0.0 ± 0.1 | 1.5 ± 0.5 | 0.1 ± 0.3 |
| Missing | 0 | 2 | 0 | 2 |
| MDS-UPDRS part 1 score | 5.4 ± 3.9 | 3.0 ± 3.1 | 4.7 ± 3.8 | 2.8 ± 2.7 |
| Missing | 1 | 1 | 0 | 0 |
| MDS-UPDRS part 2 score | 5.8 ± 4.0 | 0.5 ± 1.0 | 5.4 ± 4.5 | 0.1 ± 0.5 |
| Missing | 1 | 1 | 0 | 0 |
| MDS-UPDRS part 3 off score | 20.9 ± 8.7 | 1.1 ± 2.1 | 19.6 ± 9.3 | 1.7 ± 3.3 |
| Missing | 0 | 2 | 0 | 0 |
| MDS-UPDRS total off score | 32.2 ± 12.9 | 4.6 ± 4.4 | 29.7 ± 13.4 | 4.7 ± 5.2 |
| Missing | 1 | 2 | 1 | 0 |
| MOCA total score | 27.1 ± 2.3 | 28.2 ± 1.1 | 27.5 ± 1.8 | 28.3 ± 1.1 |
| Missing | 0 | 0 | 0 | 0 |
| ESS total score | 5.8 ± 3.4 | 5.6 ± 3.4 | 5.3 ± 3.3 | 4.3 ± 2.0 |
| Missing | 1 | 2 | 1 | 0 |
| RBDSQ total score | 5.1 ± 2.7 | 3.7 ± 2.2 | 4.6 ± 2.3 | 4.3 ± 2.3 |
| Missing | 4 | 0 | 0 | 0 |
| SCOPA-AUT total score | 12.4 ± 9.1 | 8.2 ± 7.8 | 11.1 ± 8.7 | 7.0 ± 5.4 |
| Missing | 4 | 1 | 1 | 0 |
| HVLT-R delayed recall T score | 44.3 ± 11.4 | 48.9 ± 10.9 | 47.7 ± 10.7 | 51.5 ± 10.8 |
| Missing | 0 | 0 | 0 | 0 |
| JLO adjusted total score | 12.2 ± 2.9 | 12.5 ± 2.9 | 11.4 ± 3.1 | 12.5 ± 3.2 |
| Missing | 0 | 0 | 0 | 0 |
| SDMT total T score | 45.4 ± 9.0 | 50.1 ± 10.5 | 44.1 ± 8.7 | 51.8 ± 10.7 |
| Missing | 0 | 0 | 0 | 0 |
| SBR (Caudate + Putamen) | 4.2 ± 1.1 | 7.8 ± 1.7 | 4.3 ± 1.0 | 7.4 ± 1.3 |
| Missing | 12 | 30 | 4 | 1 |
| SBR (Caudate) | 3.0 ± 0.8 | 4.5 ± 0.9 | 3.0 ± 0.8 | 4.3 ± 0.7 |
| Missing | 12 | 30 | 4 | 1 |
| SBR (Putamen) | 1.2 ± 0.4 | 3.3 ± 0.9 | 1.3 ± 0.4 | 3.0 ± 0.6 |
| Missing | 12 | 30 | 4 | 1 |
| American Indian/Alaska Native<sup>a</sup> | 4 (1.3%) | 2 (1.2%) | 0 (0.0%) | 0 (0.0%) |
| Asian<sup>a</sup> | 10 (3.2%) | 0 (0.0%) | 1 (2.6%) | 1 (6.7%) |
| Black/African American<sup>a</sup> | 7 (2.2%) | 6 (3.7%) | 0 (0.0%) | 3 (20.0%) |
| Hispanic/Latino<sup>a</sup> | 9 (2.9%) | 4 (2.5%) | 0 (0.0%) | 0 (0.0%) |
| White<sup>a</sup> | 293 (93.9%) | 151 (93.8%) | 37 (97.4%) | 11 (73.3%) |

Values are presented as means ± standard deviations or numbers (percentages).
<sup>a</sup>In the self-report of one's own race and ethnicity, multiple choices were allowed.
ESS = Epworth sleepiness scale; HVLT-R = Hopkins verbal learning test-revised; JLO = judgment of line orientation; RBDSQ = rapid-eye-movement sleep behaviour disorder screening questionnaire; SCOPA-AUT = scales for outcomes in Parkinson's disease-autonomic; SDMT = symbol digit modality test.

### Quantification of CSF proteomic signatures associated with non-genetic PD

In the derivation cohort, 149 proteins targeted by 155 SOMAmers were likely to be differentially expressed between patients with non-genetic PD and HCs (*q* ≤ 0.2) (Fig. 2A). The KEGG and Reactome pathway enrichment analyses revealed that pathways that were likely to be down-regulated in patients with non-genetic PD included those associated with the neuronal system, glutamatergic synapse, circadian entrainment, and insulin secretion (Fig. 2B). No potentially up-regulated pathways in patients with non-genetic PD were identified.

**Figure 2.**
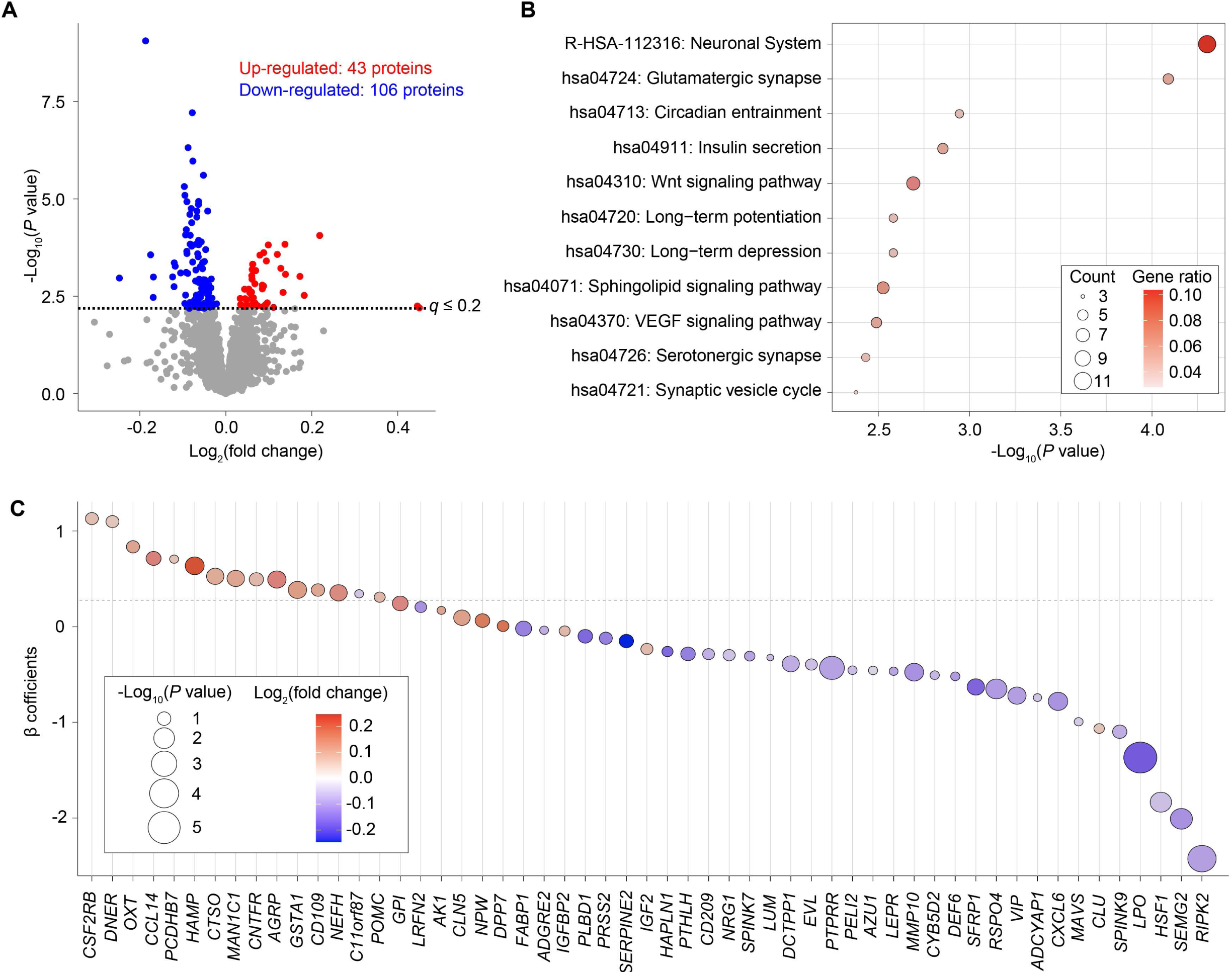
Construction of the ngPD-ProS. (**A**) Volcano plot illustrating the difference in CSF levels of each examined protein. At the *q*-value cutoff of 0.2, 149 proteins were identified as likely to be differentially expressed between patients with non-genetic PD and HCs. (**B**) KEGG and Reactome pathway enrichment analyses of proteins that were likely to be down-regulated in non-genetic PD. “Count” is the number of input proteins annotated to a pathway and “Gene ratio” is the ratio of input proteins annotated to a pathway to total proteins annotated to a pathway. (**C**) Plot depicting the values of β coefficients of each protein included in the final model used to calculate the ngPD-ProS, along with information on *P* values and log2 fold change obtained by the differentially expressed protein analysis.

A subsequent logistic regression model with LASSO selected 55 proteins (targeted by 55 SOMAmers) to construct an optimal model for computing the ngPD-ProS. The β coefficients of the 55 proteins are illustrated in Fig. 2C, and further details are presented in Supplementary Table 1. The final equation to compute the ngPD-ProS was as follows:

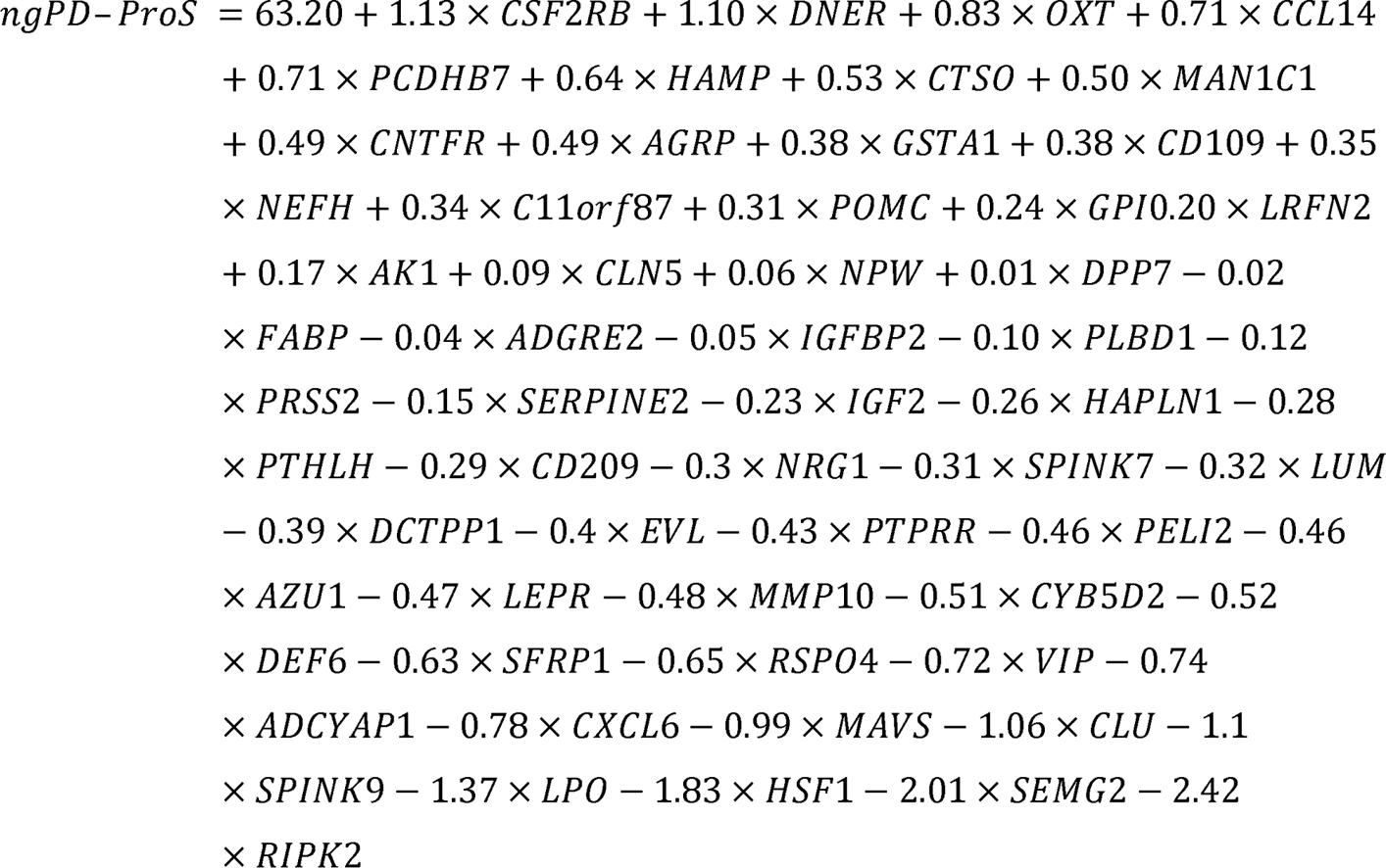

The AUC of the ngPD-ProS for distinguishing patients with non-genetic PD from HCs was 0.91 (95% CI, 0.89–0.94) in the derivation cohort (Fig. 3A). When adjusted for age and sex, the ORs for PD of participants in the middle and highest tertiles of the ngPD-ProS relative to those in the lowest tertile of the ngPD-ProS were 14.7 (95% CI, 8.6–25.9) and 86.7 (95% CI, 38.1–228.5), respectively (Fig. 3B).

**Figure 3.**
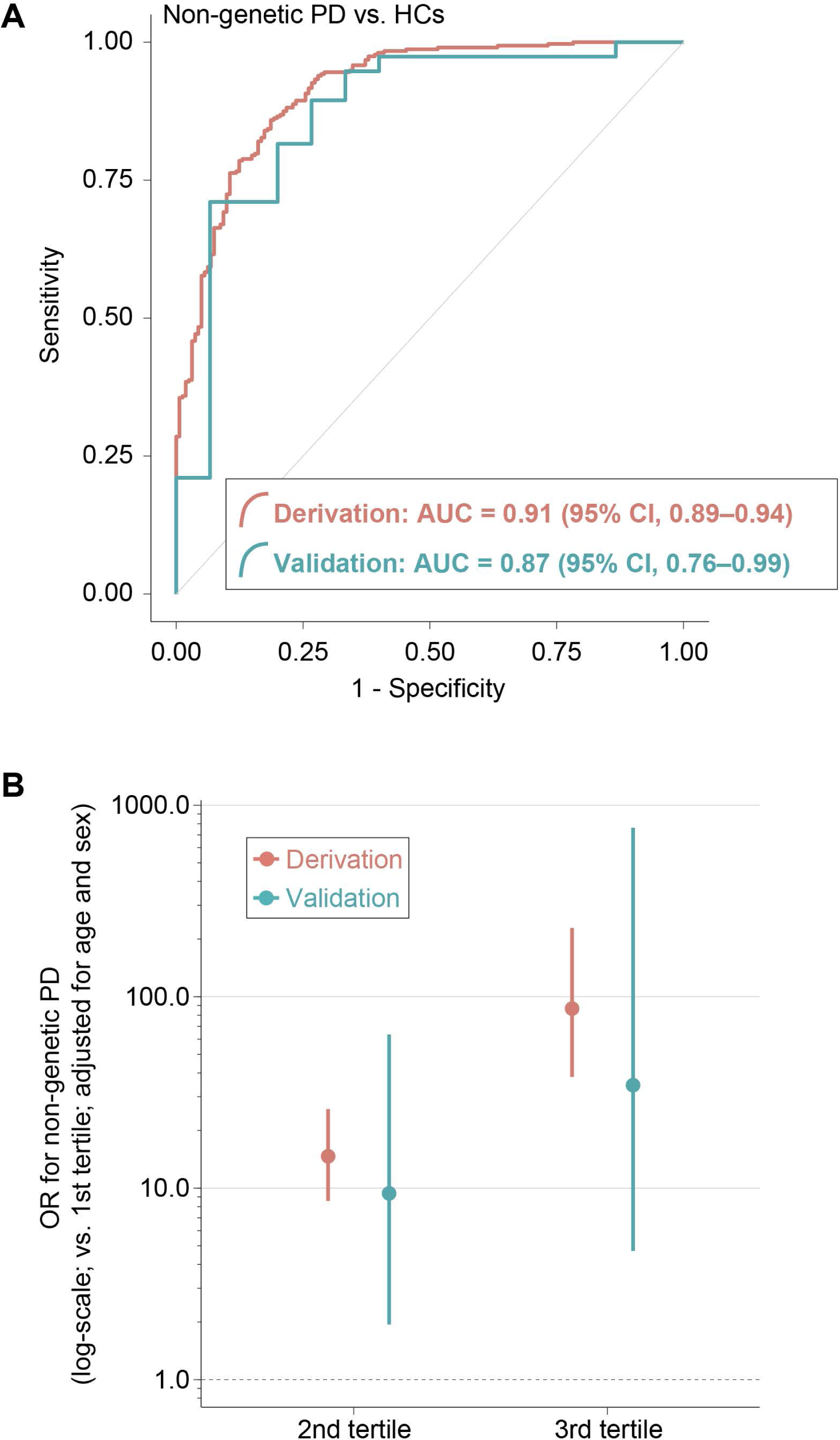
Examination of the performance of the ngPD-ProS. (**A**) Receiver operating characteristic (ROC) curves of the ngPD-ProS for the derivation and validation datasets. (**B**) Plot illustrating the ORs for non-genetic PD (adjusted for age and sex) of participants in the 2nd and 3rd tertiles of the ngPD-ProS compared with those in the lowest tertile of the ngPD-ProS. The length of lines represents 95% CI.

In the validation cohort independent of the derivation cohort, the AUC of the ngPD-ProS for distinguishing PD from HCs was 0.87 (95% CI, 0.76–0.99), which was not significantly different to the AUC in the derivation cohort (*P* = 0.51) (Fig. 3A). When adjusted for age and sex, the ORs of participants in the middle and highest tertiles of the ngPD-ProS relative to those in the lowest tertile of the ngPD-ProS were 9.4 (95% CI, 1.9–63.4) and 34.5 (95% CI, 4.7–761.9), respectively (Fig. 3B). These results collectively demonstrated that the CSF proteomic signatures associated with non-genetic PD could be accurately quantified in the form of the ngPD-ProS.

### ngPD-ProS in the genetic PD cohort

The genetic PD cohort consisted of 96 patients with *GBA1* mutations (*GBA1*-PD), 144 patients with *LRRK2* mutations (*LRRK2*-PD), 10 patients with both *GBA1* and *LRRK2* mutations (*GBA1+LRRK2*-PD), and 8 patients with *SNCA* mutations (*SNCA*-PD). Clinical characteristics of patients with genetic PD are summarized in Table 2. Details of the mutations are presented in Supplementary Table 2. The ngPD-ProS was significantly higher in patients of all genetic PD groups than in HCs (Fig. 4A). In comparison to patients with non-genetic PD, the ngPD-ProS was significantly lower in patients with *LRRK2*-PD and comparable in patients with *GBA1*-PD, *GBA1*+*LRRK2*-PD, and *SNCA*-PD. (Fig. 4A). These results indicated that the CSF proteomic signatures associated with non-genetic PD were present in patients with genetic PD, albeit to varying degrees depending on the type of mutation.

**Table 2.**
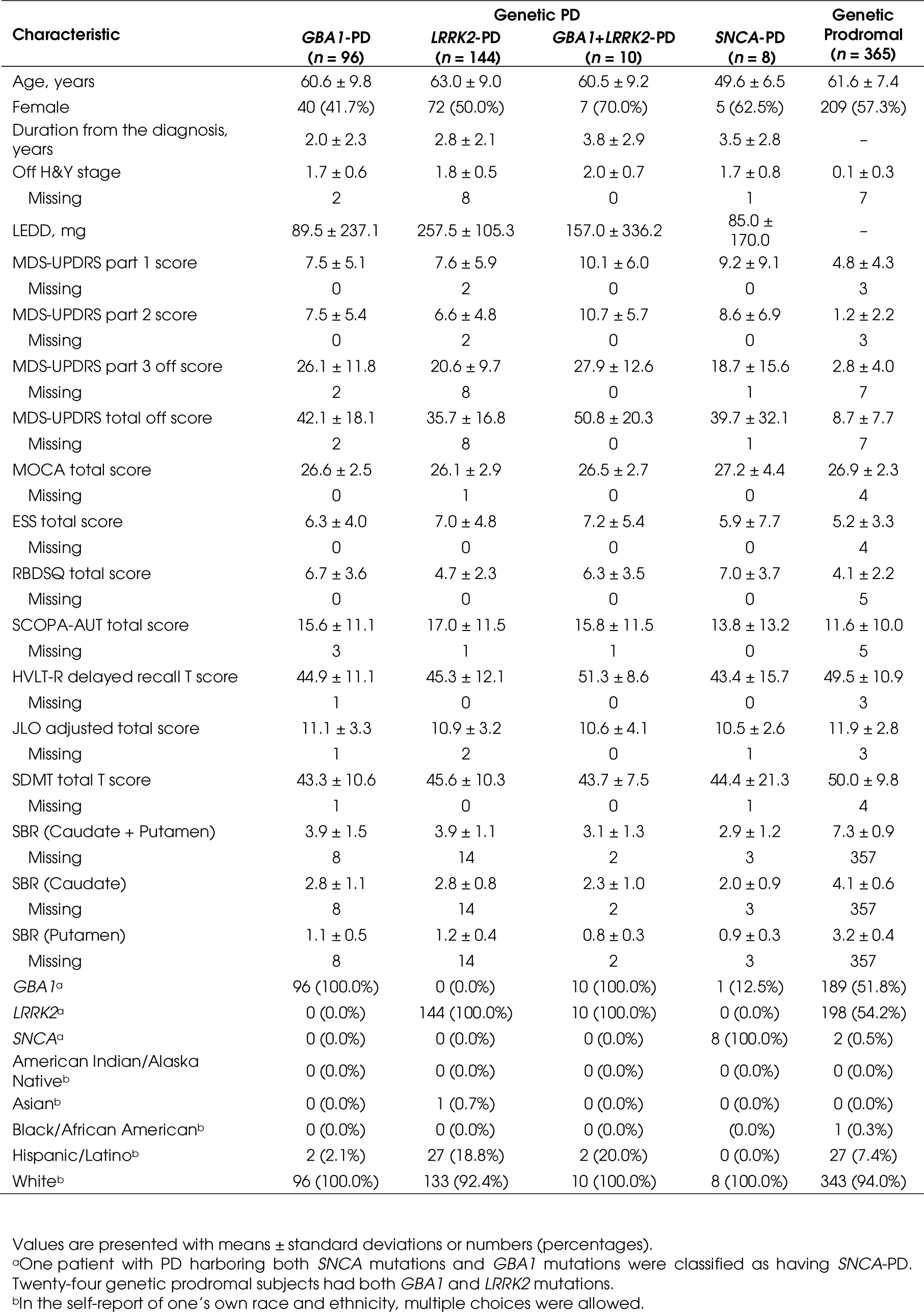
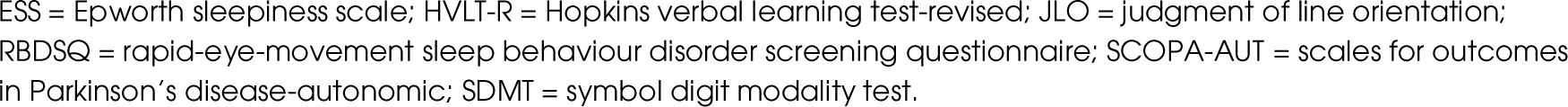
Baseline characteristics of patients with genetic PD and genetic prodromal participants included for additional analysis.

| Characteristic | Genetic PD |  |  |  | Genetic Prodromal (n = 365) |
| --- | --- | --- | --- | --- | --- |
|  | *GBA1*-PD (n = 96) | *LRRK2*-PD (n = 144) | *GBA1*+*LRRK2*-PD (n = 10) | *SNCA*-PD (n = 8) |  |
| Age, years | 60.6 ± 9.8 | 63.0 ± 9.0 | 60.5 ± 9.2 | 49.6 ± 6.5 | 61.6 ± 7.4 |
| Female | 40 (41.7%) | 72 (50.0%) | 7 (70.0%) | 5 (62.5%) | 209 (57.3%) |
| Duration from the diagnosis, years | 2.0 ± 2.3 | 2.8 ± 2.1 | 3.8 ± 2.9 | 3.5 ± 2.8 | – |
| Off H&Y stage | 1.7 ± 0.6 | 1.8 ± 0.5 | 2.0 ± 0.7 | 1.7 ± 0.8 | 0.1 ± 0.3 |
| Missing | 2 | 8 | 0 | 1 | 7 |
| LEDD, mg | 89.5 ± 237.1 | 257.5 ± 105.3 | 157.0 ± 336.2 | 85.0 ± 170.0 | – |
| MDS-UPDRS part 1 score | 7.5 ± 5.1 | 7.6 ± 5.9 | 10.1 ± 6.0 | 9.2 ± 9.1 | 4.8 ± 4.3 |
| Missing | 0 | 2 | 0 | 0 | 3 |
| MDS-UPDRS part 2 score | 7.5 ± 5.4 | 6.6 ± 4.8 | 10.7 ± 5.7 | 8.6 ± 6.9 | 1.2 ± 2.2 |
| Missing | 0 | 2 | 0 | 0 | 3 |
| MDS-UPDRS part 3 off score | 26.1 ± 11.8 | 20.6 ± 9.7 | 27.9 ± 12.6 | 18.7 ± 15.6 | 2.8 ± 4.0 |
| Missing | 2 | 8 | 0 | 1 | 7 |
| MDS-UPDRS total off score | 42.1 ± 18.1 | 35.7 ± 16.8 | 50.8 ± 20.3 | 39.7 ± 32.1 | 8.7 ± 7.7 |
| Missing | 2 | 8 | 0 | 1 | 7 |
| MOCA total score | 26.6 ± 2.5 | 26.1 ± 2.9 | 26.5 ± 2.7 | 27.2 ± 4.4 | 26.9 ± 2.3 |
| Missing | 0 | 1 | 0 | 0 | 4 |
| ESS total score | 6.3 ± 4.0 | 7.0 ± 4.8 | 7.2 ± 5.4 | 5.9 ± 7.7 | 5.2 ± 3.3 |
| Missing | 0 | 0 | 0 | 0 | 4 |
| RBDSQ total score | 6.7 ± 3.6 | 4.7 ± 2.3 | 6.3 ± 3.5 | 7.0 ± 3.7 | 4.1 ± 2.2 |
| Missing | 0 | 0 | 0 | 0 | 5 |
| SCOPA-AUT total score | 15.6 ± 11.1 | 17.0 ± 11.5 | 15.8 ± 11.5 | 13.8 ± 13.2 | 11.6 ± 10.0 |
| Missing | 3 | 1 | 1 | 0 | 5 |
| HVLT-R delayed recall T score | 44.9 ± 11.1 | 45.3 ± 12.1 | 51.3 ± 8.6 | 43.4 ± 15.7 | 49.5 ± 10.9 |
| Missing | 1 | 0 | 0 | 0 | 3 |
| JLO adjusted total score | 11.1 ± 3.3 | 10.9 ± 3.2 | 10.6 ± 4.1 | 10.5 ± 2.6 | 11.9 ± 2.8 |
| Missing | 1 | 2 | 0 | 1 | 3 |
| SDMT total T score | 43.3 ± 10.6 | 45.6 ± 10.3 | 43.7 ± 7.5 | 44.4 ± 21.3 | 50.0 ± 9.8 |
| Missing | 1 | 0 | 0 | 1 | 4 |
| SBR (Caudate + Putamen) | 3.9 ± 1.5 | 3.9 ± 1.1 | 3.1 ± 1.3 | 2.9 ± 1.2 | 7.3 ± 0.9 |
| Missing | 8 | 14 | 2 | 3 | 357 |
| SBR (Caudate) | 2.8 ± 1.1 | 2.8 ± 0.8 | 2.3 ± 1.0 | 2.0 ± 0.9 | 4.1 ± 0.6 |
| Missing | 8 | 14 | 2 | 3 | 357 |
| SBR (Putamen) | 1.1 ± 0.5 | 1.2 ± 0.4 | 0.8 ± 0.3 | 0.9 ± 0.3 | 3.2 ± 0.4 |
| Missing | 8 | 14 | 2 | 3 | 357 |
| *GBA1*<sup>a</sup> | 96 (100.0%) | 0 (0.0%) | 10 (100.0%) | 1 (12.5%) | 189 (51.8%) |
| *LRRK2*<sup>a</sup> | 0 (0.0%) | 144 (100.0%) | 10 (100.0%) | 0 (0.0%) | 198 (54.2%) |
| *SNCA*<sup>a</sup> | 0 (0.0%) | 0 (0.0%) | 0 (0.0%) | 8 (100.0%) | 2 (0.5%) |
| American Indian/Alaska Native<sup>b</sup> | 0 (0.0%) | 0 (0.0%) | 0 (0.0%) | 0 (0.0%) | 0 (0.0%) |
| Asian<sup>b</sup> | 0 (0.0%) | 1 (0.7%) | 0 (0.0%) | 0 (0.0%) | 0 (0.0%) |
| Black/African American<sup>b</sup> | 0 (0.0%) | 0 (0.0%) | 0 (0.0%) | 0 (0.0%) | 1 (0.3%) |
| Hispanic/Latino<sup>b</sup> | 2 (2.1%) | 27 (18.8%) | 2 (20.0%) | 0 (0.0%) | 27 (7.4%) |
| White<sup>b</sup> | 96 (100.0%) | 133 (92.4%) | 10 (100.0%) | 8 (100.0%) | 343 (94.0%) |

Values are presented with means ± standard deviations or numbers (percentages).
<sup>a</sup>One patient with PD harboring both *SNCA* mutations and *GBA1* mutations were classified as having *SNCA*-PD. Twenty-four genetic prodromal subjects had both *GBA1* and *LRRK2* mutations.
<sup>b</sup>In the self-report of one's own race and ethnicity, multiple choices were allowed.
ESS = Epworth sleepiness scale; HVLt-R = Hopkins verbal learning test-revised; JLO = judgment of line orientation; RBDSQ = rapid-eye-movement sleep behaviour disorder screening questionnaire; SCOPA-AUT = scales for outcomes in Parkinson's disease-autonomic; SDMT = symbol digit modality test.

**Figure 4.**
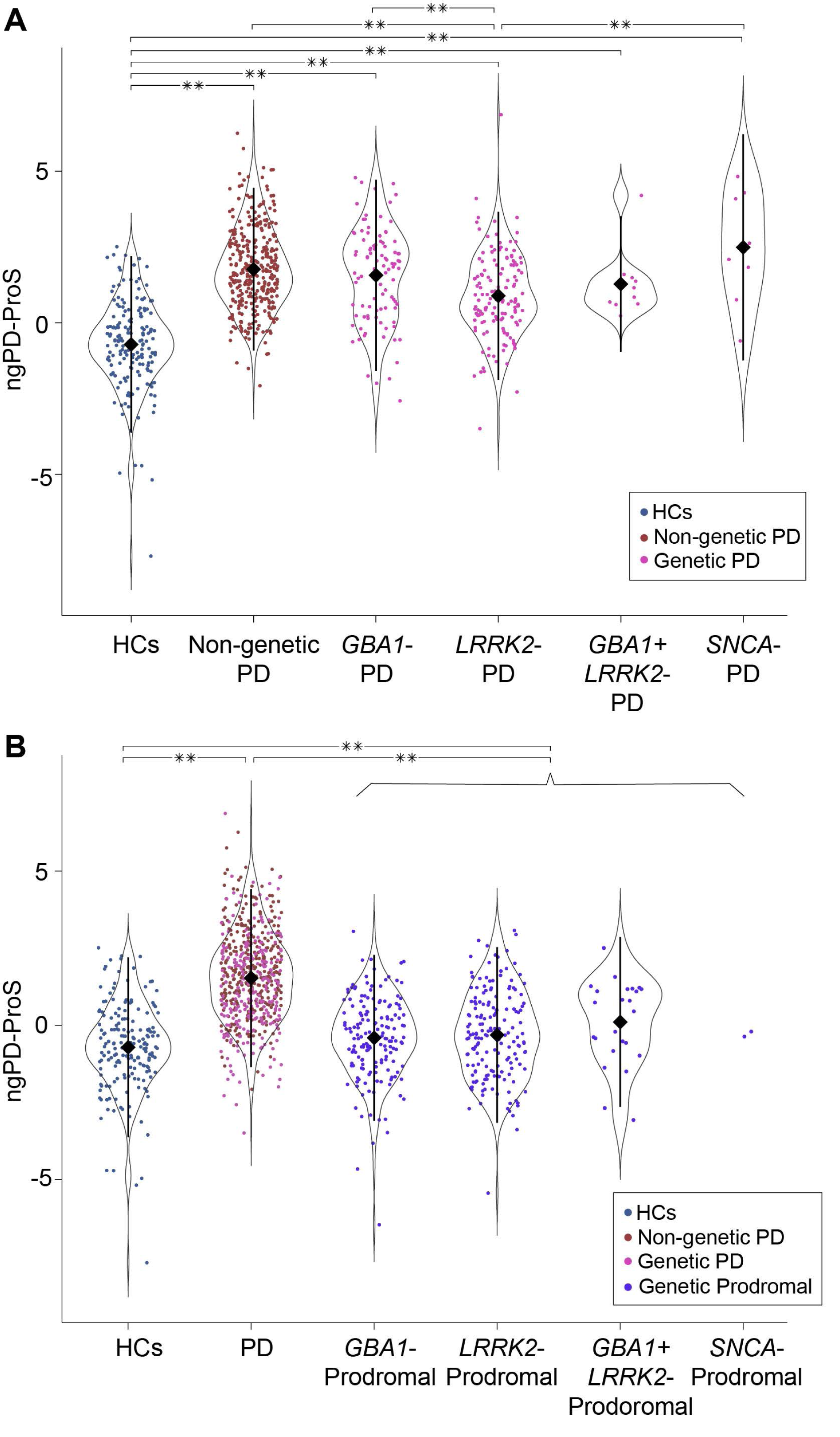
Examination of the ngPD-ProS in the genetic PD and genetic prodromal cohorts. (**A**) Dot and volcano plots illustrating the ngPD-ProS of HCs, non-genetic PD, and genetic PD. All genetic PD groups exhibited significantly higher ngPD-ProS than HCs, albeit to varying degrees depending on the type of mutation (-0.7 ± 1.5 [HCs] vs. 1.8 ± 1.3 [non-genetic PD] vs. 1.6 ± 1.6 [*GBA1*-PD] vs. 0.9 ± 1.4 [*LRRK2*-PD] vs. 1.3 ± 1.1 [*GBA1*+*LRRK2*-PD] vs. 2.5 ± 1.9 [*SNCA*-PD]). Diamonds and error bars represent the mean and standard deviation, respectively. \*\**q* ≤ 0.01. (**B**) Dot and volcano plots illustrating the ngPD-ProS of HCs, patients with PD, and genetic prodromal participants. The ngPD-ProS of genetic prodromal participants was significantly higher than that of HCs and significantly lower than that of patients with PD (-0.7 ± 1.5 [HCs] vs. 1.5 ± 1.4 [PD] vs. -0.3 ± 1.4 [genetic prodromal]). Since there were only 2 genetic prodromal participants with *SNCA* mutations, individual data were plotted for genetic prodromal participants with *SNCA* mutations. Diamonds and error bars represent the mean and standard deviation, respectively. \*\**q* ≤ 0.01.

### ngPD-ProS in the genetic prodromal cohort

The genetic prodromal cohort consisted of 165 participants with *GBA1* mutations, 174 participants with *LRRK2* mutations, 24 participants with both *GBA1* and *LRRK2* mutations, and 2 participants with *SNCA* mutations. Clinical characteristics of participants in the genetic prodromal cohort are summarized in Table 2. Details of the mutations are presented in Supplementary Table 3. The ngPD-ProS of genetic prodromal participants was significantly higher than that of HCs and remained significantly lower than that of patients with PD (Fig. 4B). The difference between the types of mutations did not significantly differ in genetic prodromal participants (all *q* ≥ 0.49). These results suggested that the emergence of non-genetic PD-associated CSF proteomic signatures preceded the onset of PD.

### Correlation analysis of the ngPD-ProS with baseline factors

Serial correlation analyses of baseline factors of patients with PD included in all cohorts (*n* = 608) revealed that even after adjusting for age, sex, LEDD, and the presence of *GBA1*, *LRRK2*, and/or *SNCA* mutations, the ngPD-ProS was significantly correlated with several baseline clinical parameters (Fig. 5A); among these, the strongest correlations were found with the SBR of DAT imaging (caudate, *r_par_* = -0.19; caudate plus putamen, *r_par_* = -018; both *q* < 0.001). In a subset of patients with PD for whom data were available for all CSF biomarkers examined in this study (*n* = 198) (Supplementary Table 4), the ngPD-ProS was also significantly correlated with several baseline CSF biomarkers including amyloid-β 1–42 (*r_par_*= -0.27, *q* < 0.001) and α-synuclein (*r_par_* = -0.28, *q* < 0.001) (Fig. 5B). The results remained relatively unchanged when patients with PD were sub-grouped into patients with non-genetic PD and those with genetic PD (Fig. 5A and B). However, no significant associations were observed between baseline factors and the ngPD-ProS in HCs (Fig. 5A and B). These results collectively implied that the ngPD-ProS could be used to quantify various pathologies relevant to PD symptoms.

**Figure 5.**
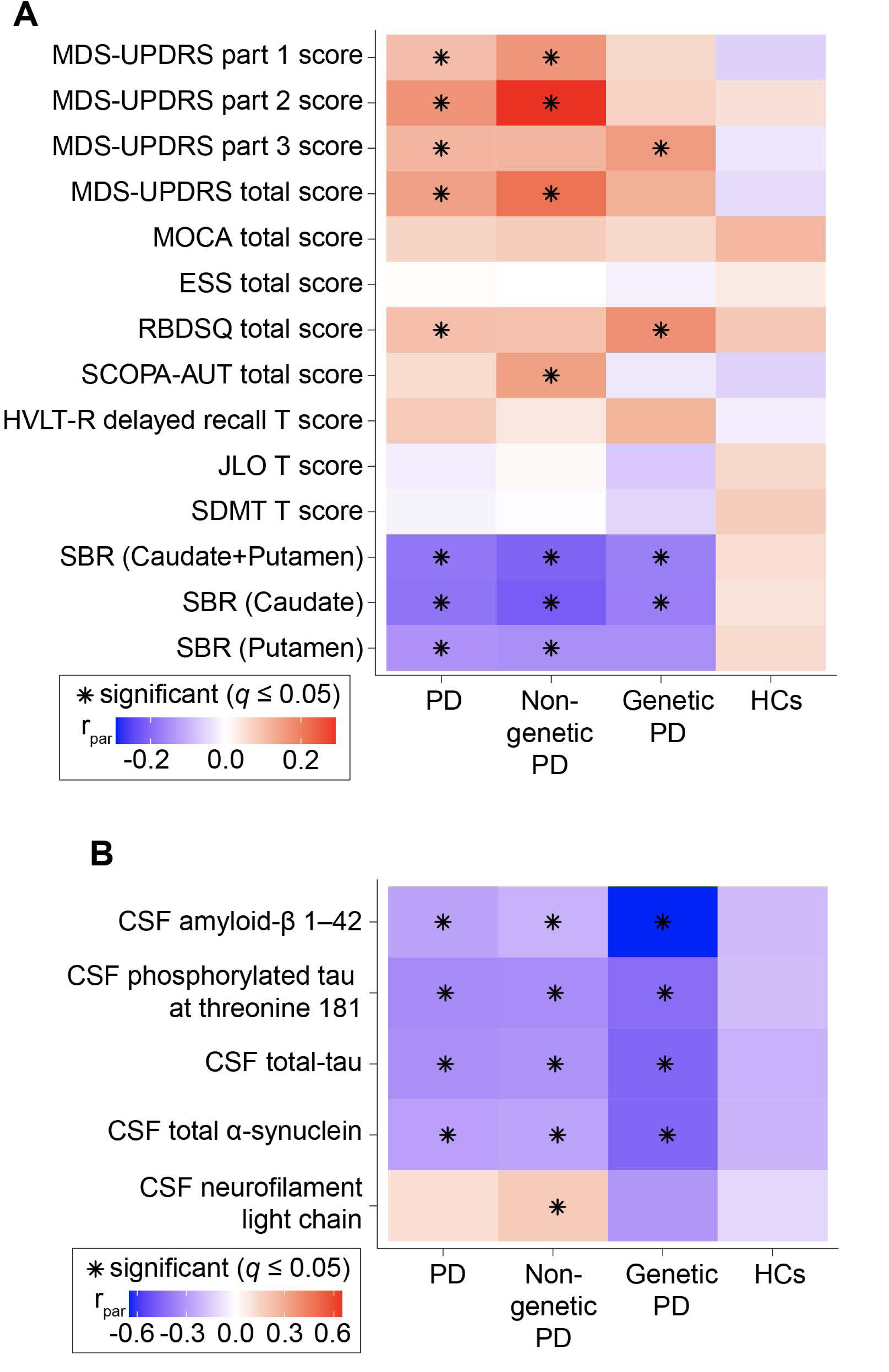
Correlation analysis of the ngPD-ProS with baseline factors. (**A**) Heatmap showing the values of *r_par_* between the ngPD-ProS and baseline clinical factors. \**q* ≤ 0.05. (**B**) Heatmap showing the values of *r_par_* between the ngPD-ProS and baseline CSF biomarkers. \**q* ≤ 0.05. ESS = Epworth sleepiness scale; HVLT-R = Hopkins verbal learning test-revised; JLO = judgment of line orientation; RBDSQ = rapid-eye-movement sleep behavior disorder screening questionnaire; SCOPA-AUT = scales for outcomes in Parkinson’s disease-autonomic; SDMT = symbol digit modality test.

### Impact of the ngPD-ProS on the development of important clinical millstones

As longitudinal clinical milestones, MCI, dementia, and H&Y stages III and IV were selected. Details of the number of events and loss to follow-up are summarized in Supplementary Table 5. Preliminary univariate Cox regression analysis revealed that a significantly increased risk of faster progression became evident after the ngPD-ProS exceeded its 90th percentile value (Supplementary Fig. 2). Therefore, the remaining analyses were performed by comparing patients with PD within the highest ngPD-ProS decile with the remaining patients. Serial Kaplan–Meier analyses demonstrated that patients with PD within the highest ngPD-ProS decile had a significantly faster progression to MCI (χ^2^ = 16.8; *q* < 0.001) (Fig. 6A), dementia (χ^2^ = 20.4; *q* < 0.001) (Fig. 6B), and H&Y stage IV (χ^2^ = 16.0; *q* < 0.001) (Fig. 6C). The results were similar even after adjusting for age, sex, baseline LEDD, the presence of *GBA1*, *LRRK2*, and/or *SNCA* mutations, baseline MDS-UPDRS part III score, baseline MOCA total score, and baseline SBR value of DAT imaging via the Cox proportional hazard model (MCI, adjusted HR [aHR] = 2.0 [95% CI, 1.2–3.2; *q* = 0.01]; dementia, aHR = 2.4 [95% CI, 1.2–4.8; *q* = 0.02]; H&Y stage IV, aHR = 2.6 [95% CI, 1.2–5.5; *q* = 0.02]) (Fig. 6D). Sensitivity analysis revealed the same results (Supplementary Table 6).

**Figure 6.**
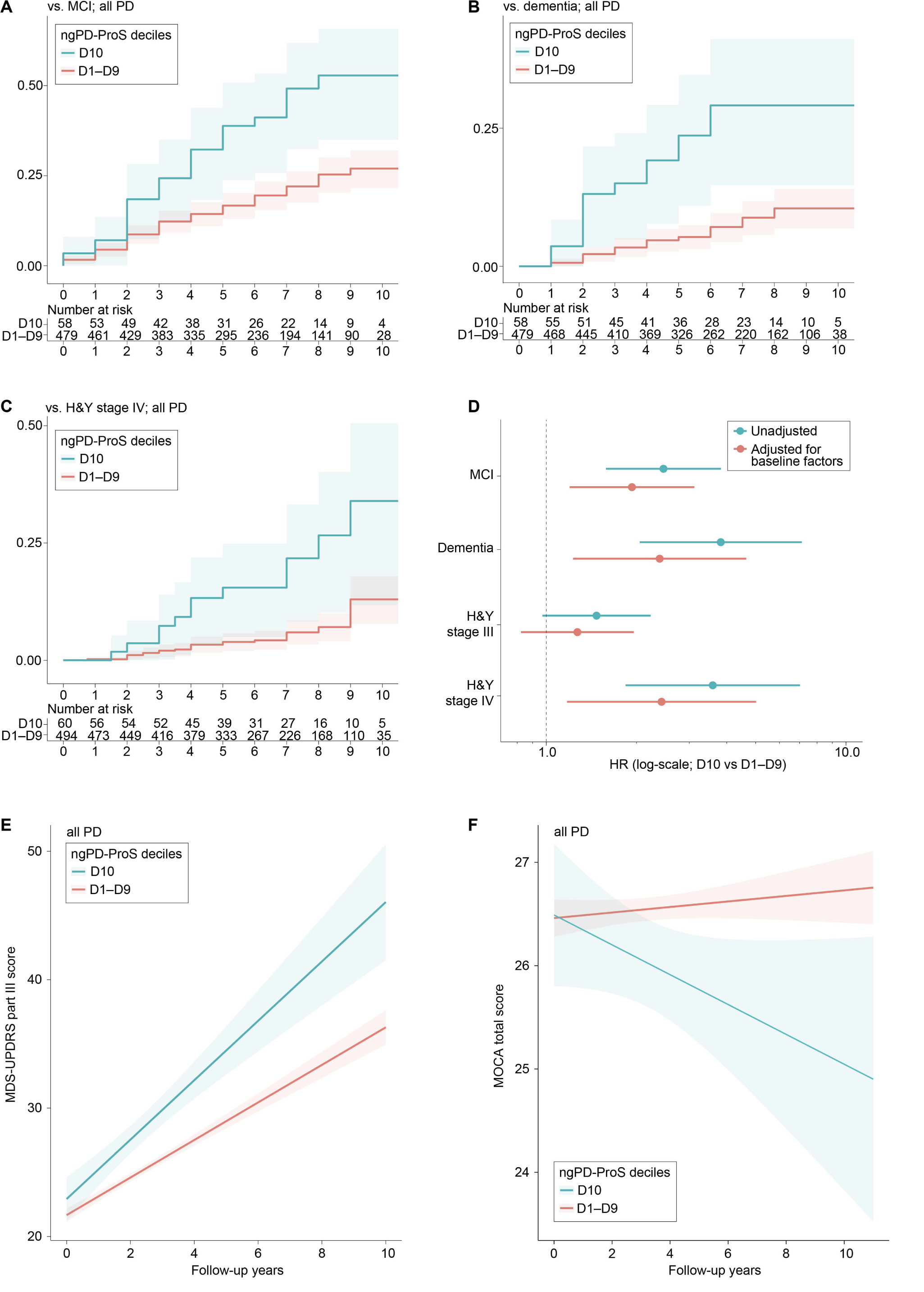
Longitudinal analysis of the impact of the ngPD-ProS on the subsequent clinical course of PD. (**A**) Kaplan−Meier curve illustrating the difference in the rate of MCI development between patients with PD in the highest decile (D10) of the ngPD-ProS compared with those in the 1st to 9th deciles (D1–D9) of the ngPD-ProS. Shaded area represents 95% CI. (**B**) Kaplan−Meier curve illustrating the difference in the rate of dementia development between patients with PD in D10 of the ngPD-ProS compared with those in D1– D9 of the ngPD-ProS. Shaded area represents 95% CI. (**C**) Kaplan−Meier curve illustrating the difference in the rate of progression to H&Y stage IV between patients with PD in D10 of the ngPD-ProS compared with those in D1–D9 of the ngPD-ProS. Shaded area represents 95% CI. (**D**) Forest plot depicting HRs (non-adjusted and adjusted for age; sex; baseline LEDD; the presence of *GBA1*, *LRRK2*, and/or *SNCA* mutations; baseline MDS-UPDRS part III and MOCA total scores; and SBR value of DAT imaging) of patients with PD in D10 of the ngPD-ProS compared with those in D1–D9 of the ngPD-ProS. The length of lines represents 95% CI. (**E**) Linear regression lines showing the difference in temporal changes in motor symptoms quantified by the MDS-UPDRS part III score between patients with PD in D10 of the ngPD-ProS compared with those in D1–D9 of the ngPD-ProS. Shaded area represents 95% CI. (**F**) Linear regression lines showing the difference in temporal changes in cognitive symptoms quantified by the MOCA total score between patients with PD in D10 of the ngPD-ProS compared with those in D1–D9 of the ngPD-ProS. Shaded area represents 95% CI.

### Interaction of the ngPD-ProS with subsequent motor and cognitive decline

Finally, the interaction of the ngPD-ProS with longitudinal changes in MDS-UPDRS part III score and MOCA total score was examined via the multivariate linear mixed effects model adjusted for age, sex, LEDD, the presence of *GBA1*, *LRRK2*, and/or *SNCA* mutations, baseline MDS-UPDRS part III score, baseline MOCA total score, and baseline SBR value of DAT imaging. It was found that patients with PD within the highest ngPD-ProS decile had a significantly faster progression of motor symptoms (fixed-effects coefficients of the interaction term [β_interaction_] = 0.89 [95% CI, 0.14–1.6]; t = 2.3; *q* = 0.03) (Fig. 6E) and cognitive symptoms (β_interaction_ = -0.46 [95% CI, -0.64 to -0.28]; t = -5.0; *q* < 0.001) (Fig. 6F). These results collectively indicated that the subsequent clinical course varied greatly depending on the degree to which non-genetic PD-associated CSF proteomic signatures were present in each patient.

## Discussion

In the current study, via a combination of untargeted high-throughput proteomics and machine-learning, we derived and validated a 55-protein-based ngPD-ProS that accurately captured CSF proteomic signatures associated with non-genetic PD. Based on this score, we identified that non-genetic PD-associated CSF proteomic signatures were also present in patients with genetic PD to varying degrees depending on the type of mutation and appeared from the prodromal phase. Further, we have shown that non-genetic PD-associated CSF proteomic signatures were correlated with several baseline clinical parameters and CSF biomarkers, and were robustly linked to subsequent motor and cognitive decline in PD, regardless of genetic or non-genetic PD. In conjunction with the findings of an article related to ours,^33^ this study provides the most comprehensive proteomic profiling of PD to date. Furthermore, to our knowledge, this is the first study to successfully develop a measure that accurately captures non-genetic PD-associated CSF proteomic signatures and to demonstrate its robust association with the progression of motor and cognitive symptoms of PD, collectively indicating that the CSF proteome encodes information important for both the onset and progression of PD.

This study confirms the utility of extracting information as signatures from multidimensional proteomic data. Even though the differences in the levels of each protein may be modest, when combined as signatures, non-genetic PD and HCs could be reliably distinguished (AUC = 0.91 [derivation cohort] and 0.87 [validation cohort]). This is also true in the field of genomics, where the polygenic risk score (PRS), which is calculated by summing the effects of multiple risk variants that have modest effects alone, holds great promise in explaining disease susceptibility.^66–68^ Crucially, the ngPD-ProS constructed from 312 patients with non-genetic PD and 161 HCs in this study demonstrated better diagnostic accuracy compared to the PRS constructed in a recent large-scale genome-wide association study involving around 37,700 patients with PD and 1,400,000 HCs (AUC = 0.64 [derivation cohort] and 0.69 [validation cohort]).^68^ While genomics is undoubtedly important and have played a major role in elucidating the pathophysiology of PD,^69–74^ one of its limitations is that the information encoded in the germline genome is “static” and does reflect the dynamic state of the brain. Although future replication studies are warranted, our results clearly highlighted the importance of non-genetic information captured by high-throughput proteomics for encapsulating the pathological features associated with PD.

We believe that the strength of the ngPD-ProS is that it can contribute not only to diagnosis of PD but also to stratification of both cognitive and motor decline in patients with PD, despite its solely proteomics-dependent derivation. Recent pioneering studies have reported that combining multiple pieces of information, including disease-associated protein levels, the presence of α-synuclein seeds, genomics, transcriptomics, and clinical information, can improve the accuracy of diagnosis and/or stratification.^75–79^ However, previous studies have rarely succeeded in improving all aspects of diagnosis, motor symptom stratification, and cognitive symptom stratification.^80^ It has also been reported that the polygenic susceptibility that drives the onset of PD and the progression of symptoms may not be the same.^81–83^

Therefore, we are initially surprised that the ngPD-ProS can not only distinguish PD from HCs but also have a significant impact on the subsequent disease course. Admittedly, it is difficult to precisely explain the biological mechanism underlying our results, but we conjecture that the ngPD-ProS might be a unique measure that can sensitively reflect overall changes in the brains of patients with PD, including the burden of abnormal protein deposition, which are crucial for the clinical course of PD.^84, 85^ This is because1) the ngPD-ProS should reflect down-regulation of the neuronal system in general and pathways that have been reported to be associated with PD (Fig. 2B),^86–89^ 2) the ngPD-ProS was significantly correlated with several CSF biomarkers (although proteomics cannot measure either disease-associated forms or “seeds” of α-synuclein), 3) the ngPD-ProS was slightly increased even in the prodromal participants, and 4) the ngPD-ProS predicted the progression of cognitive and motor symptoms. This remains speculative and, therefore, should be tested in future pathological studies.

Finally, it should be mentioned that in an article related to ours,^33^ it was found that the most differentially expressed proteins in *GBA1*-PD, *LRRK2*-PD, and non-genetic PD differed from each other. Therefore, at first, we did not expect to observe that CSF proteomic signatures associated with non-genetic PD also existed in patients with *GBA1*-PD, *LRRK2*-PD, and *SNCA*-PD. However, we believe that this is very reasonable given that although the primary pathophysiological mechanism driving the disease might not be the same among these PD subtypes, they have much in common at least in terms of pathological changes (i.e., dopaminergic neuronal loss, accompanied by α-synuclein aggregates in most cases).^90, 91^ Accordingly, the fact that *LRRK2*-PD often lacks α-synuclein pathology supports our observation that the CSF proteomic signatures associated with non-genetic PD were least shared with *LRRK2*-PD.^92^ Importantly, from a genomic perspective, it has also been shown that polygenic burden of PD is associated with the development of PD not only in non-genetic PD but also in *GBA1*-PD and *LRRK2*-PD.^93–95^

The strengths of our study were as follows: 1) patients with non-genetic PD that were used to construct the ngPD-ProS were relatively large in the sample size and relatively uniform since they were drug-naïve and within 2-years of diagnosis; and 2) the inclusion of patients with genetic PD and genetic prodromal participants as well as comprehensive baseline and follow-up data enabled the assessment of the ngPD-ProS from multiple perspectives. However, several limitations should also be mentioned. The main limitation of our study is the overrepresentation of participants with European ancestry in the PPMI study.^96, 97^ Therefore, the generalizability of the current results to other populations warrants further studies. In addition, the need for replication in other large-scale cohorts cannot be stressed enough.^98^ Other limitations include insufficient data in the PPMI database to analyze the ability of the ngPD-ProS to discriminate PD from atypical parkinsonism, longitudinal changes in the ngPD-ProS, and the impact of the ngPD-ProS on future PD development in genetic prodromal participants.

In conclusion, this large-scale CSF proteomic study demonstrated that CSF proteomic signatures associated with non-genetic PD can be quantified using the novel ngPD-ProS. Furthermore, through multi-step and comprehensive analyses using this score, our study confirmed the great potential of high-throughput proteomics for elucidating the pathophysiology and developing biomarkers for PD, and moreover, highlighted the great potential of CSF proteomic signatures as a surrogate indicator of the dynamic brain state of patients with PD.

## Data Availability

All data used in this study are available for certified investigators in the PPMI database. The R scripts used in this study will be made publicly available upon acceptance.

## Abbreviations

AUC: area under the receiver operating characteristic curve;
CI: confidence interval;
CSF: cerebrospinal fluid;
DAT: dopamine transporter;
HCs: healthy controls;
MCI: mild cognitive impairment;
HR: hazard ratio;
LEDD: levodopa-equivalent daily dose;
MDS-UPDRS: Movement disorder society-sponsored revision of the unified Parkinson’s disease rating scale;
MOCA: Montreal cognitive assessment;
OR: odds ratio;
PD: Parkinson’s disease;
ngPD-ProS: non-genetic Parkinson’s disease-associated proteomic score;
PPMI: Parkinson’s Progression Markers Initiative;
PRS: polygenic risk score;
*r_par_*: Pearson’s partial correlation coefficient;
SBR: specific binding ratio;
SOMAmers: slow offrate modified aptamers.

## Acknowledgements

The authors thank Dr. Takahiro Kamada, who died in January 2019, for inspiring them to conduct this study, and they dedicate this article to his memory. PPMI – a public-private partnership – is funded by the Michael J. Fox Foundation for Parkinson’s Research funding partners 4D Pharma, Abbvie, Acurex Therapeutics, Allergan, Amathus Therapeutics, ASAP, Avid Radiopharmaceuticals, Bial Biotech, Biogen, BioLegend, Bristol-Myers Squibb, Calico, Celgene, Dacapo Brain Science, Denali, The Edmond J. Safra Foundation, GE Healthcare, Genentech, GlaxoSmithKline, Golub Capital, Handl Therapeutics, Insitro, Janssen Neuroscience, Lilly, Lundbeck, Merck, Meso Scale Discovery, Neurocrine Biosciences, Pfizer, Piramal, Prevail, Roche, Sanofi Genzyme, Servier, Takeda, Teva, UCB, Verily, and Voyager.

