## Supplementary Material for "Changes in the cerebrospinal fluid proteome precede and stratify the course of Parkinson’s Disease"

**Supplementary Figures**

**Supplementary Figure 1 Study flowchart.**

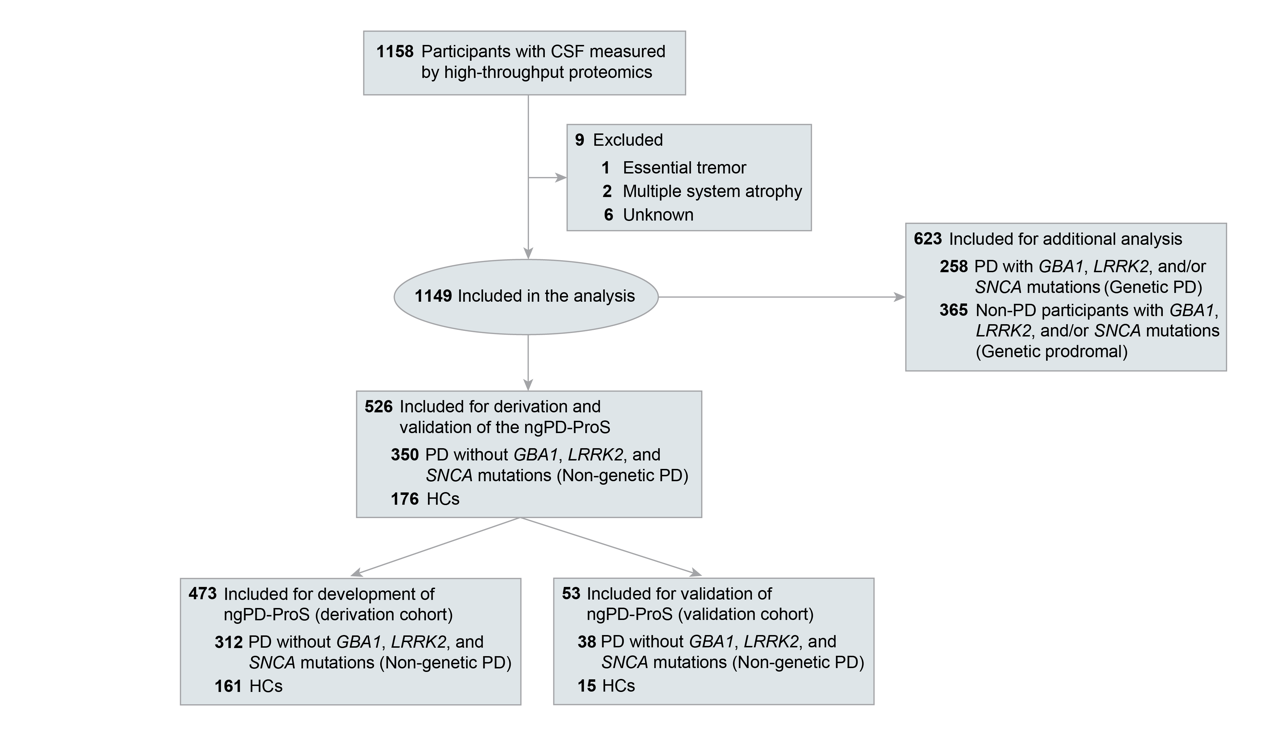

**Supplementary Figure 2 Dose-response relationship between the ngPD-ProS and risk of developing clinical milestones.** Note that the risk of developing clinical milestones appeared to increase in patients with PD in the highest decile (D10) of the ngPD-ProS compared with those in the lowest decile (D1) of the ngPD-ProS.

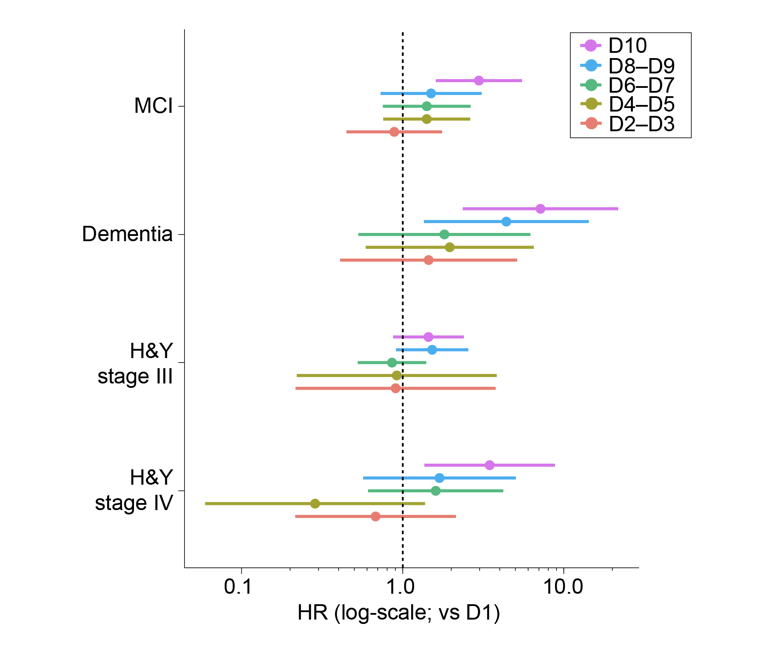

**Supplementary Tables**

**Supplementary Table 1 Details of 55 proteins included in the tuned model to calculate the ngPD-ProS**

| **SOMAmers ID** | **Target gene** | **Derivation cohort** | | **Validation cohort** | | **Beta coefficient** |
| --- | --- | --- | --- | --- | --- | --- |
|  |  | **log2(FC)** | **OR (95%CI)** | **log2(FC)** | **OR (95%CI)** |  |
| 11137-43_3 | CSF2RB | 0.05 | 1.49 (1.21–1.86) | 0.01 | 1.07 (0.58–2.15) | 1.13 |
| 9769-48_3 | DNER | 0.04 | 1.02 (0.84–1.24) | -0.09 | 0.76 (0.37–1.42) | 1.10 |
| 8356-88_3 | OXT | 0.09 | 1.38 (1.13–1.70) | 0.10 | 1.51 (0.80–3.07) | 0.83 |
| 2900-53_3 | CCL14 | 0.14 | 1.69 (1.36–2.11) | 0.13 | 1.35 (0.71–2.75) | 0.71 |
| 8071-114_3 | PCDHB7 | 0.05 | 1.41 (1.15–1.75) | 0.00 | 1.01 (0.54–1.92) | 0.71 |
| 3504-58_2 | HAMP | 0.22 | 1.74 (1.40–2.18) | 0.75 | 6.29 (2.46–22.1) | 0.64 |
| 9264-11_3 | CTSO | 0.08 | 1.22 (1.01–1.49) | -0.05 | 0.85 (0.44–1.56) | 0.53 |
| 13427-66_3 | MAN1C1 | 0.09 | 1.29 (1.06–1.58) | -0.07 | 0.78 (0.40–1.50) | 0.50 |
| 2813-11_2 | AGRP | 0.14 | 1.42 (1.15–1.76) | -0.24 | 0.60 (0.30–1.11) | 0.49 |
| 2711-6_2 | CNTFR | 0.06 | 1.01 (0.83–1.23) | -0.09 | 0.75 (0.37–1.42) | 0.49 |
| 12446-49_3 | GSTA1 | 0.10 | 1.55 (1.23–2.00) | 0.06 | 1.32 (0.71–2.73) | 0.38 |
| 3290-50_2 | CD109 | 0.09 | 1.27 (1.04–1.55) | 0.03 | 1.12 (0.56–2.24) | 0.38 |
| 9900-36_3 | NEFH | 0.12 | 1.72 (1.31–2.28) | -0.02 | 0.95 (0.47–2.05) | 0.35 |
| 11116-16_3 | C11orf87 | -0.04 | 0.72 (0.59–0.88) | -0.11 | 0.58 (0.29–1.09) | 0.34 |
| 2558-51_3 | POMC | 0.06 | 1.33 (1.09–1.64) | -0.04 | 0.84 (0.46–1.59) | 0.31 |
| 4272-46_2 | GPI | 0.13 | 1.21 (0.99–1.49) | 0.11 | 1.57 (0.70–3.80) | 0.24 |
| 7200-4_3 | LRFN2 | -0.09 | 0.66 (0.52–0.81) | -0.21 | 0.66 (0.32–1.24) | 0.20 |
| 5012-67_1 | AK1 | 0.09 | 1.15 (0.94–1.42) | 0.14 | 1.70 (0.84–3.94) | 0.17 |
| 8874-53_3 | CLN5 | 0.09 | 1.57 (1.25–1.99) | -0.09 | 0.71 (0.37–1.33) | 0.09 |
| 9986-14_3 | NPW | 0.17 | 1.47 (1.20–1.80) | 0.25 | 1.42 (0.77–2.71) | 0.06 |
| 8346-9_3 | DPP7 | 0.18 | 1.24 (1.03–1.50) | 0.04 | 1.06 (0.56–1.99) | 0.01 |
| 11516-7_3 | FABP1 | -0.12 | 0.98 (0.78–1.23) | -0.02 | 0.94 (0.48–1.89) | -0.02 |
| 4546-27_3 | ADGRE2 | -0.06 | 0.77 (0.63–0.93) | 0.03 | 1.18 (0.64–2.23) | -0.04 |
| 8819-3_3 | IGFBP2 | 0.06 | 1.50 (1.22–1.85) | 0.13 | 1.92 (0.99–4.06) | -0.05 |
| 6315-58_3 | PLBD1 | -0.17 | 0.86 (0.70–1.05) | -0.22 | 0.72 (0.35–1.35) | -0.10 |
| 5034-79_1 | PRSS2 | -0.12 | 0.92 (0.75–1.12) | 0.05 | 1.15 (0.60–2.29) | -0.12 |
| 3217-74_2 | SERPINE2 | -0.25 | 0.77 (0.63–0.94) | 0.00 | 1 (0.54–1.87) | -0.15 |
| 6973-111_4 | IGF2 | 0.06 | 1.51 (1.23–1.88) | 0.16 | 2.81 (1.31–7.16) | -0.23 |
| 3196-6_2 | HAPLN1 | -0.17 | 0.69 (0.57–0.84) | -0.23 | 0.69 (0.34–1.3) | -0.26 |
| 2962-50_2 | PTHLH | -0.12 | 0.75 (0.61–0.91) | -0.20 | 0.53 (0.25–1.01) | -0.28 |
| 3029-52_2 | CD209 | -0.06 | 0.73 (0.59–0.89) | -0.01 | 0.94 (0.49–1.78) | -0.29 |
| 9178-30_3 | NRG1 | -0.04 | 0.72 (0.57–0.88) | 0.03 | 1.29 (0.70–2.40) | -0.30 |
| 10974-20_3 | SPINK7 | -0.07 | 0.93 (0.75–1.15) | -0.12 | 0.55 (0.26–1.08) | -0.31 |
| 13114-50_3 | LUM | -0.05 | 0.83 (0.67–1.01) | -0.02 | 0.86 (0.44–1.68) | -0.32 |
| 4314-12_2 | DCTPP1 | -0.06 | 0.87 (0.71–1.07) | -0.17 | 0.48 (0.21–0.97) | -0.39 |
| 11656-110_3 | EVL | -0.04 | 0.75 (0.61–0.92) | -0.09 | 0.66 (0.32–1.26) | -0.40 |
| 6361-49_3 | PTPRR | -0.08 | 0.61 (0.48–0.76) | -0.16 | 0.61 (0.28–1.17) | -0.43 |
| 12702-13_3 | PELI2 | -0.05 | 0.74 (0.59–0.90) | -0.08 | 0.57 (0.26–1.08) | -0.46 |
| 14713-46_3 | AZU1 | -0.02 | 0.92 (0.76–1.12) | 0.01 | 1.18 (0.59–2.44) | -0.46 |
| 5400-52_3 | LEPR | -0.07 | 0.78 (0.63–0.95) | 0.00 | 1.01 (0.53–1.85) | -0.47 |
| 8479-4_3 | MMP10 | -0.09 | 0.76 (0.62–0.92) | -0.15 | 0.49 (0.22–0.94) | -0.48 |
| 10924-258_3 | CYB5D2 | -0.05 | 0.74 (0.60–0.91) | -0.12 | 0.57 (0.27–1.10) | -0.51 |
| 14090-23_3 | DEF6 | -0.08 | 0.78 (0.64–0.94) | 0.05 | 1.31 (0.66–3.20) | -0.52 |
| 3221-54_1 | SFRP1 | -0.17 | 0.66 (0.53–0.81) | -0.20 | 0.61 (0.29–1.16) | -0.63 |
| 8464-31_3 | RSPO4 | -0.08 | 0.65 (0.52–0.80) | -0.12 | 0.53 (0.26–1.01) | -0.65 |
| 3522-57_1 | VIP | -0.08 | 0.63 (0.51–0.78) | -0.13 | 0.67 (0.34–1.26) | -0.72 |
| 8285-64_3 | ADCYAP1 | -0.04 | 0.70 (0.57–0.85) | -0.03 | 0.71 (0.37–1.29) | -0.74 |
| 3495-15_2 | CXCL6 | -0.09 | 0.88 (0.72–1.07) | -0.03 | 0.91 (0.49–1.73) | -0.78 |
| 8783-216_3 | MAVS | -0.03 | 0.88 (0.72–1.06) | -0.01 | 0.95 (0.51–1.86) | -0.99 |
| 4542-24_2 | CLU | 0.05 | 1.42 (1.16–1.76) | 0.03 | 1.19 (0.63–2.32) | -1.06 |
| 8042-88_3 | SPINK9 | -0.06 | 0.78 (0.64–0.94) | -0.09 | 0.46 (0.18–1.04) | -1.10 |
| 4801-13_3 | LPO | -0.19 | 0.54 (0.42–0.68) | -0.30 | 0.30 (0.11–0.63) | -1.37 |
| 11616-9_3 | HSF1 | -0.04 | 0.72 (0.59–0.88) | -0.05 | 0.71 (0.35–1.37) | -1.83 |
| 6373-54_3 | SEMG2 | -0.10 | 0.68 (0.54–0.84) | -0.10 | 0.62 (0.32–1.14) | -2.01 |
| 8993-151_3 | RIPK2 | -0.08 | 0.53 (0.43–0.66) | -0.14 | 0.37 (0.16–0.74) | -2.42 |

Values are presented with 95%CIs.

FC = fold change.

**Supplementary Table 2 Details of the mutations of patients with genetic PD**

| **Mutation type** | **PD with *GBA1* (*n* = 107), *LRRK2* (*n* = 154), and *SNCA* (*n* = 8) mutations^a^** |
| --- | --- |
| ***GBA1* mutation** | |
| 84GG: 1allele | 3 (2.8%) |
| IVS2+1G>A: 1allele | 1 (0.9%) |
| R120W: 1allele | 1 (0.9%) |
| S173*: 1allele | 0 (0.0%) |
| E326K: 1allele | 23 (21.5%) |
| T369M: 1allele | 11 (10.3%) |
| N370S: 1allele | 57 (53.3%) |
| N370S: 2allele | 4 (3.7%) |
| E427K: 1allele | 1 (0.9%) |
| L483P: 1allele | 3 (2.8%) |
| R502C: 1allele | 1 (0.9%) |
| R535H: 1allele | 1 (0.9%) |
| ***LRRK2* mutation** | |
| N1437H: 1allele | 1 (0.6%) |
| R1441C: 1allele | 1 (0.6%) |
| R1441G: 1allele | 15 (9.7%) |
| G2019S: 1allele | 134 (87.0%) |
| G2019S: 2allele | 3 (1.9%) |
| I2020T: 1allele | 0 (0.0%) |
| ***SNCA* mutation** | |
| A53T: 1 allele | 8 (100.0%) |

Values are presented as numbers (percentages).

^a^There were 10 patients with PD harbouring both *GBA1* and *LRRK2* mutations and one patient with PD harbouring *GBA1* and *SNCA* mutations.

PD = Parkinson’s disease.

**Supplementary Table 3 Details of the mutations of genetic prodromal participants**

| **Mutation type** | **Prodromal subjects with GBA1 (*n* = 189), LRRK2 (*n* = 198), and SNCA (*n* = 2) mutations^a^** |
| --- | --- |
| ***GBA1* mutation** | |
| 84GG: 1allele | 0 (0.0%) |
| IVS2+1G>A: 1allele | 0 (0.0%) |
| R120W: 1allele | 2 (1.1%) |
| S173*: 1allele | 0 (0.0%) |
| E326K: 1allele | 9 (4.8%) |
| T369M: 1allele | 4 (2.1%) |
| N370S: 1allele | 158 (83.6%) |
| N370S: 2allele | 12 (6.3%) |
| E427K: 1allele | 0 (0.0%) |
| L483P: 1allele | 3 (1.6%) |
| R502C: 1allele | 0 (0.0%) |
| R535H: 1allele | 0 (0.0%) |
| ***LRRK2* mutation** | |
| N1437H: 1allele | 0 (0.0%) |
| R1441C: 1allele | 0 (0.0%) |
| R1441G: 1allele | 16 (8.1%) |
| G2019S: 1allele | 182 (91.9%) |
| I2020T: 1allele | 0 (0.0%) |
| ***SNCA* mutation** | |
| A53T: 1 allele | 2 (100.0%) |

Values are presented as numbers (percentages).

^a^There were 24 genetic prodromal participants with both *GBA1* and *LRRK2* mutations.

**Supplementary Table 4 Baseline characteristics of patients with PD and HCs for whom CSF biomarker data were available**

| **Characteristic** | **PD**  **(*n* = 198)** | **HCs**  **(*n* = 95)** | ***q* value^a^** |
| --- | --- | --- | --- |
| Age, years | 61.1 ± 10.0 | 61.4 ± 11.0 | 0.70 |
| Female | 132 (66.7%) | 61 (64.2%) | 0.70 |
| Duration from the diagnosis, years | 0.6 ± 0.7 | – | – |
| LEDD, mg | 0.0 ± 0.0 | – | – |
| CSF Aβ_1-42_, pg/mL | 886.4 ± 400.7 | 1012.2 ± 513.0 | 0.18 |
| CSF p-tau, pg/mL | 14.1 ± 5.3 | 17.1 ± 9.0 | 0.03 |
| CSF t-tau, pg/mL | 163.3 ± 57.1 | 191.8 ± 81.7 | 0.03 |
| CSF α-synuclein, pg/mL | 101.6 ± 48.7 | 128.4 ± 54.9 | < 0.01 |
| CSF NfL, pg/mL | 102.3 ± 58.6 | 97.8 ± 56.3 | 0.70 |
| *GBA1* | 26 (13.1%) | 0 (0.0%) | < 0.01 |
| *LRRK2* | 4 (2.1%) | 0 (0.0%) | 0.55 |
| *SNCA* | 0 (0.0%) | 0 (0.0%) | – |
| American Indian/Alaska Native | 2 (1.0%) | 0 (0.0%) | > 0.9 |
| Asian | 8 (4.0%) | 1 (1.1%) | 0.55 |
| Black/African American | 2 (1.0%) | 5 (5.3%) | 0.14 |
| Hispanic/Latino | 6 (3.0%) | 1 (1.1%) | 0.55 |
| White | 188 (94.9%) | 88 (92.6%) | 0.55 |

Values are presented as means ± standard deviations or numbers (percentages).

^a^Two-sample t-test and Fisher's exact test were used, as appropriate, and multiple testing was adjusted using the Benjamini & Hochberg method.

Aβ_1-42_ = amyloid-β 1 to 42; NfL = neurofilament light chain; p-tau = phosphorylated tau at threonine 181; t-tau = total-tau.

**Supplementary Table 5 Details of the number of events and loss to follow-up in the longitudinal survival analysis**

| Years | **Outcome: MCI** | | **Outcome: Dementia** | | **Outcome: H&Y stage III** | | **Outcome: H&Y stage IV** | |
| --- | --- | --- | --- | --- | --- | --- | --- | --- |
|  | **Lost**  (n = 419) | **Event**  (*n* = 118) | **Lost**  (*n* = 489) | **Event**  (*n* = 48) | **Lost**  (*n* = 375) | **Event**  (*n* = 179) | **Lost**  (*n* = 512) | **Event**  (*n* = 42) |
| 0 | 3 (0.7%) | 10 (8.5%) | 3 (0.6%) | 0 (0.0%) | 6 (1.6%) | 16 (8.9%) | 6 (1.2%) | 0 (0%) |
| 0.25 | – | – | – | – | 4 (1.1%) | 3 (1.7%) | 4 (0.8%) | 0 (0%) |
| 0.5 | 10 (2.4%) | 0 (0.0%) | 11 (2.3%) | 0 (0.0%) | 11 (2.9%) | 9 (5.0%) | 13 (2.5%) | 0 (0%) |
| 0.75 | – | – | – | – | 1 (0.3%) | 7 (3.9%) | 1 (0.2%) | 1 (2.4%) |
| 1 | 21 (5.0%) | 15 (12.7%) | 22 (4.5%) | 5 (10.4%) | 12 (3.2%) | 12 (6.7%) | 13 (2.5%) | 0 (0%) |
| 1.5 | – | – | – | – | 12 (3.2%) | 9 (5.0%) | 12 (2.3%) | 1 (2.4%) |
| 2 | 28 (6.7%) | 25 (21.2%) | 29 (5.9%) | 12 (25.0%) | 12 (3.2%) | 16 (8.9%) | 16 (3.1%) | 5 (11.9%) |
| 2.5 | – | – | – | – | 9 (2.4%) | 9 (5.0%) | 12 (2.3%) | 2 (4.8%) |
| 3 | 34 (8.1%) | 18 (15.3%) | 39 (8.0%) | 6 (12.5%) | 15 (4.0%) | 11 (6.1%) | 20 (3.9%) | 4 (9.5%) |
| 3.5 | – | – | – | – | 9 (2.4%) | 10 (5.6%) | 18 (3.5%) | 2 (4.8%) |
| 4 | 35 (8.4%) | 12 (10.2%) | 41 (8.4%) | 7 (14.6%) | 19 (5.1%) | 11 (6.1%) | 29 (5.7%) | 6 (14.3%) |
| 4.5 | – | – | – | – | 11 (2.9%) | 11 (6.1%) | 17 (3.3%) | 0 (0.0%) |
| 5 | 53 (12.6%) | 11 (9.3%) | 68 (13.9%) | 4 (8.3%) | 42 (11.2%) | 10 (5.6%) | 71 (13.9%) | 3 (7.1%) |
| 6 | 37 (8.8%) | 9 (7.6%) | 40 (8.2%) | 7 (14.6%) | 37 (9.9%) | 16 (8.9%) | 44 (8.6%) | 1 (2.4%) |
| 7 | 52 (12.4%) | 9 (7.6%) | 63 (12.9%) | 4 (8.3%) | 43 (11.5%) | 10 (5.6%) | 63 (12.3%) | 6 (14.3%) |
| 8 | 49 (11.6%) | 7 (5.9%) | 57 (11.7%) | 3 (6.2%) | 51 (13.6%) | 9 (5.0%) | 61 (11.9%) | 3 (7.1%) |
| 9 | 65 (15.6%) | 2 (1.7%) | 73 (15.0%) | 0 (0.0%) | 51 (13.6%) | 8 (4.5%) | 72 (14.1%) | 8 (19.0%) |
| 10 | 29 (6.9%) | 0 (0.0%) | 39 (8.0%) | 0 (0.0%) | 27 (7.2%) | 2 (1.1%) | 37 (7.2%) | 0 (0.0%) |
| 11 | 3 (0.7%) | 0 (0.0%) | 4 (0.8%) | 0 (0.0%) | 3 (0.8%) | 0 (0.0%) | 3 (0.6%) | 0 (0.0%) |

Values are presented as numbers (percentages).

**Supplementary Table 6 Results of the sensitivity analysis for the longitudinal survival analysis.**

| **Characteristic** | **Hazard ratio (95%CI), unadjusted** | **Hazard ratio (95%CI), adjusted** |
| --- | --- | --- |
| Outcome: MCI | 2.6 (1.6–4.2) | 2.0 (1.2–3.4) |
| Outcome: Dementia | 5.0 (2.5–10.0) | 3.3 (1.6–7.6) |
| Outcome: H&Y stage III | 1.4 (0.86–2.3) | 1.4 (0.83–2.3) |
| Outcome: H&Y stage IV | 4.2 (1.8–9.8) | 3.1 (1.2–8.0) |

Values are presented with 95%CIs.
